# Deleterious heteroplasmic mitochondrial mutations increase risk of overall and cancer-specific mortality

**DOI:** 10.1101/2022.09.20.22280151

**Authors:** Stephanie L Battle, Yun Soo Hong, Wen Shi, Daniela Puiu, Vamsee Pillalamarri, Nathan Pankratz, Nicole J Lake, Monkol Lek, Eliseo Guallar, Dan E Arking

**Author notes:** Corresponding author. Correspondence and requests for materials should be addressed to Dan E. Arking. Dan E. Arking, McKusick-Nathans Institute, Department of Genetic Medicine, Johns Hopkins University School of Medicine, 733 N. Broadway, Miller Research Building, Room 459, Baltimore, MD 21205. These authors contributed equally (Stephanie L Battle, Yun Soo Hong).

## Abstract

Mitochondria are involved in energetic, biosynthetic, and homeostatic processes in eukaryotic cells. Mitochondria carry their own circular genome and disruption of the quantity or quality of mitochondrial genome is associated with various aging-related diseases^1–3^. Unlike the nuclear genome, mitochondrial DNA (mtDNA) can be present at 1,000s to 10,000s copies in somatic cells and variants may exist in a state of heteroplasmy, where only a fraction of the DNA molecules harbor a particular variant. We used MitoHPC, a bioinformatics pipeline, to accurately quantify mtDNA heteroplasmy from whole genome sequencing data in 194,871 participants in the UK Biobank. We found that the presence of heteroplasmy is associated with an increased risk of all-cause mortality (adjusted hazard ratio [aHR] 1.50-fold; 95% confidence interval [CI] 1.14, 1.98, when comparing participants with 4 or more heteroplasmies to those without any heteroplasmy). In addition, we functionally characterized mtDNA single nucleotide variants (SNVs) using a novel constraint-based score, Mitochondrial local constraint (MLC) score sum (MSS), which demonstrated that SNVs at highly constrained sites were strongly associated with all-cause mortality (aHR for a 1-unit increase in MSS 1.28; 95% CI 1.20, 1.37) and cancer-related mortality (aHR 1.36; 95% CI 1.24,1.49), particularly lung and breast cancers, lymphoma, and leukemia. MSS was also associated with prevalence and incidence of lung cancer, lymphoma, and leukemia. Moreover, among individuals with prevalent leukemia, high MSS was strongly associated with leukemia mortality (adjusted HR 4.03; 95% CI 1.34, 12.11). These results indicate that mitochondria may have a functional role in certain cancers and mitochondrial heteroplasmic SNVs have the potential to serve as a prognostic markers for cancer incidence and outcome, especially for leukemia.

## Main text

Mitochondria are the main energy production organelle of the cell and are also involved in numerous cellular processes including calcium homeostasis, apoptosis, fatty acid oxidation, and the generation of metabolic intermediates^4^. Mitochondria contain their own circular chromosome (mtDNA) that is present in 1,000s to 10,000s copies per somatic cell^4^ and is replicated independently of the nuclear genome. Its lack of protective histones, limited DNA repair system, and proximity to reactive oxygen species (ROS) producing reactions result in the acquisition of mutations in a fraction of mtDNA molecules within a cell or tissue, a state known as heteroplasmy. Heteroplasmic single nucleotide variants (SNVs) are responsible for mitochondrial disorders when present at high allele frequencies^4^ and most pathogenic mtDNA SNVs are only observed in a heteroplasmic, as opposed to homoplasmic, state^1^.

Mitochondrial heteroplasmy is common, with low level heteroplasmy observed in whole-genome sequencing (WGS) studies in up to 40-45% of samples^3,5,6^. Approximately 30% of heteroplasmies observed in any given individual are maternally inherited^7^. The effect of accumulating somatic mtDNA mutations contributes to mitochondrial dysfunction, with the most severe effects observed in tissues with the highest energy demands^8^, and the role of mitochondrial dysfunction in longevity, cancer, and degenerative diseases has been well-established^2,3^. Better understanding of the effect of the presence and level of mitochondrial heteroplasmy on common or rare diseases will provide insight into the molecular physiology of disease development and progression. Large population-based cohorts, like the UK Biobank (UKB), are useful tools to tackle this problem, particularly for evaluating the association of heteroplasmy with common diseases.

We developed a bioinformatics pipeline, MitoHPC, to accurately measure mtDNA SNVs in large WGS datasets^9^. MitoHPC’s key feature is that it constructs a consensus mitochondrial sequence for each individual, which allows for more accurate read mapping and for measuring heteroplasmy against an individual’s unique mitochondrial genome. MitoHPC performs two iterations of variant calling, first to identify major alleles, or homoplasmies, which are used to construct the consensus mitochondrial sequence, and second to call heteroplasmic variants. On simulated data, MitoHPC accurately identified all heteroplasmic SNV variants without false-positives or false-negatives. MitoHPC is also built to handle large datasets of 100,000s of samples, making this an ideal pipeline for our study.

Here we present an analysis of mtDNA heteroplasmy measured in 194,871 individuals from the UK Biobank, a large cohort of men and women aged 40-69 years recruited in 2006-2010 with uniformly collected health information and biological materials^10^. We characterize mtDNA SNVs based on their genetic features and investigate the relationship between SNVs at highly constrained sites and risk of all-cause and cause-specific mortality. We find disease phenotypes that are associated with mtDNA heteroplasmic SNVs and propose the use of mitochondrial heteroplasmic SNVs detected in the blood as a risk marker for hematological cancers.

### Heteroplasmic SNV Characteristics

After quality control procedures (**Extended Data Fig. 1** and **Extended Data Fig. 2**), we identified a total of 74,369 heteroplasmic SNVs at a 5% variant allele fraction (VAF) in the UKB with 59,414 (30.5%) out of 194,871 participants having at least 1 heteroplasmy (**Table 1, Fig. 1**). This includes 11,611 unique variant alleles occurring at 10,164 (61.8%) out of 16,443 possible mtDNA base positions (polyC homopolymer regions are excluded due to challenges in sequencing accurately). A plurality of heteroplasmic SNVs are seen in only a single individual (4,265/11,611, 36.7%) and the vast majority are extremely rare, with 89% (10,306/11,611) seen in ≤10 participants (**Table 2**).

**Table 1:** Number of heteroplasmic SNVs per participant.

| Heteroplasmy Count | Number of Participants (%) |
| --- | --- |
| 0 | 135,457 (69.5%) |
| 1 | 47,008 (24.1%) |
| 2 | 10,273 (5.3%) |
| 3 | 1,785 (0.92%) |
| 4 | 280 (0.14%) |
| 5 | 68 (0.03%) |

**Table 2:** Frequency of heteroplasmic SNVs in the population.

| Variant Allele Counts | Number of Variants |
| --- | --- |
| singletons | 4,265 |
| 2-10 | 6,041 |
| 11-100 | 1,258 |
| 101-1000 | 43 |
| 1000+ | 4 |

In this table we summarize the number of times that each SNV appears in the study population. For example, there were 4,265 heteroplasmic SNVs that appeared in only 1 participant.

**Fig. 1:**
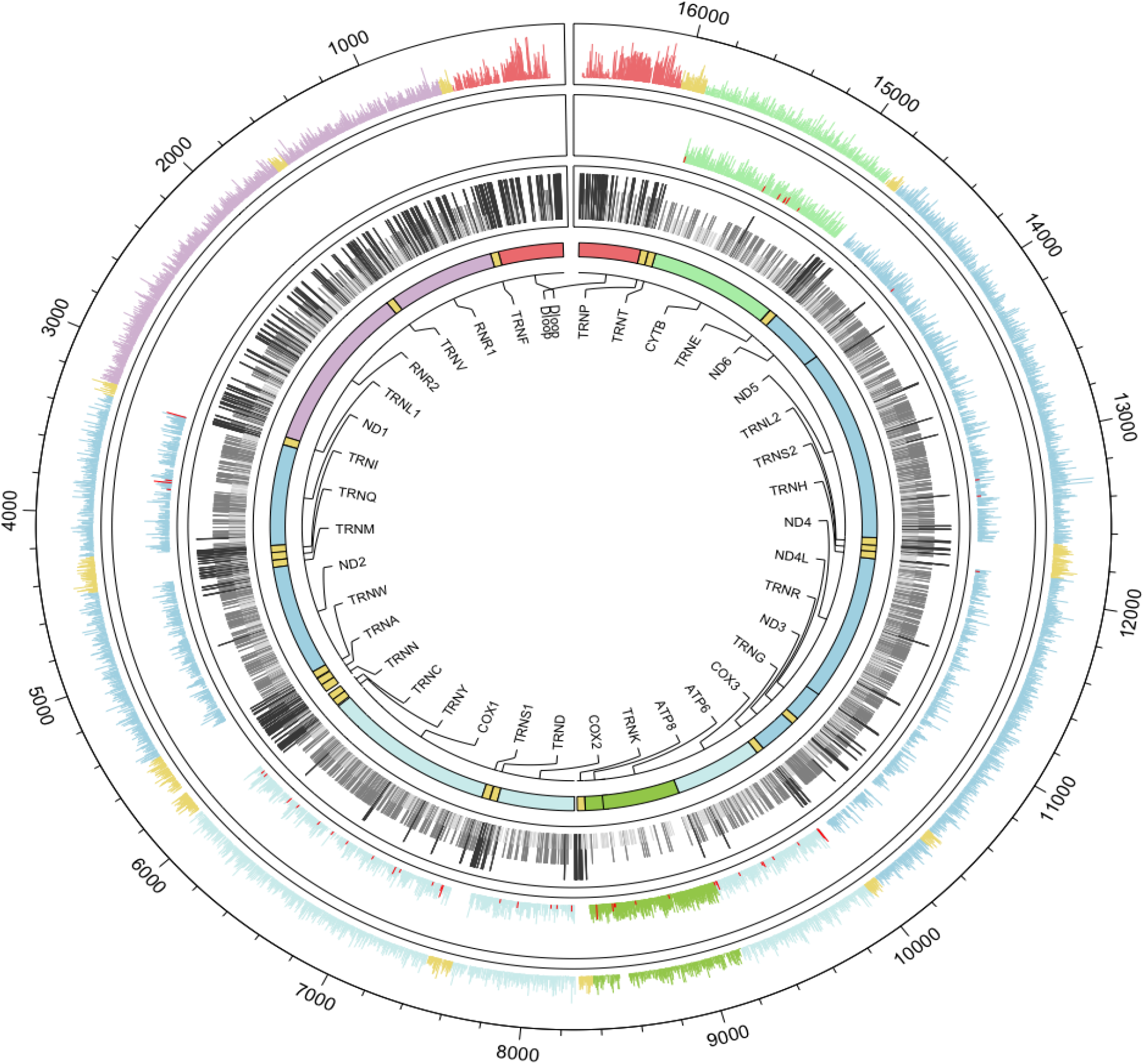
Single nucleotide variants (SNVs) and invariant sites across mitochondrial DNA. Circos plot tracks from outer (Track 1) to inner (Track 4): Track 1 is all synonymous and noncoding (tRNA, rRNA and D-loop) heteroplasmic sites. Track 2 is all nonsynonymous and nonsense heteroplasmic SNVs, where nonsense heteroplasmic SNVs are colored in red. The Y-axis of Tracks 1 and 2 is the log(number of participants with a heteroplasmy + 1), scaled from 0 to 9. Track 3 is positions with no heteroplasmy. Three or more adjacent null positions are colored light grey, 2 adjacent positions are colored medium grey, and singlets are colored dark grey. The height of Track 3 bars is scaled by color, light grey is the lowest, followed by medium grey, then dark grey. Innermost track (Track 4) is gene annotations. Genes are colored similarly by complex or by gene type.

Alleles present in 95% or more of the mtDNA WGS reads within a participant were counted as homoplasmic SNVs. A total of 4,540,614 homoplasmic SNVs were identified, with 385 individuals having no homoplasmic variants (i.e.,variants that completely match the rCRS reference mtDNA genome). Homoplasmic SNVs occurred at 7,929 unique base positions, and at 6773 (85.4%) of those sites we also observed a heteroplasmic allele. Overall, there were 14,294 unique variants found at 11,320/16,569 mtDNA sites (68.3%) (**Fig. 2a**), with 49.2% of SNVs seen as both heteroplasmic and homoplasmic, 32.0% heteroplasmic only, and 18.8% homoplasmic only (**Fig. 2b**). Thus, while most heteroplasmies are extremely rare, the majority (~60%) are also found as homoplasmies.

**Fig. 2:**
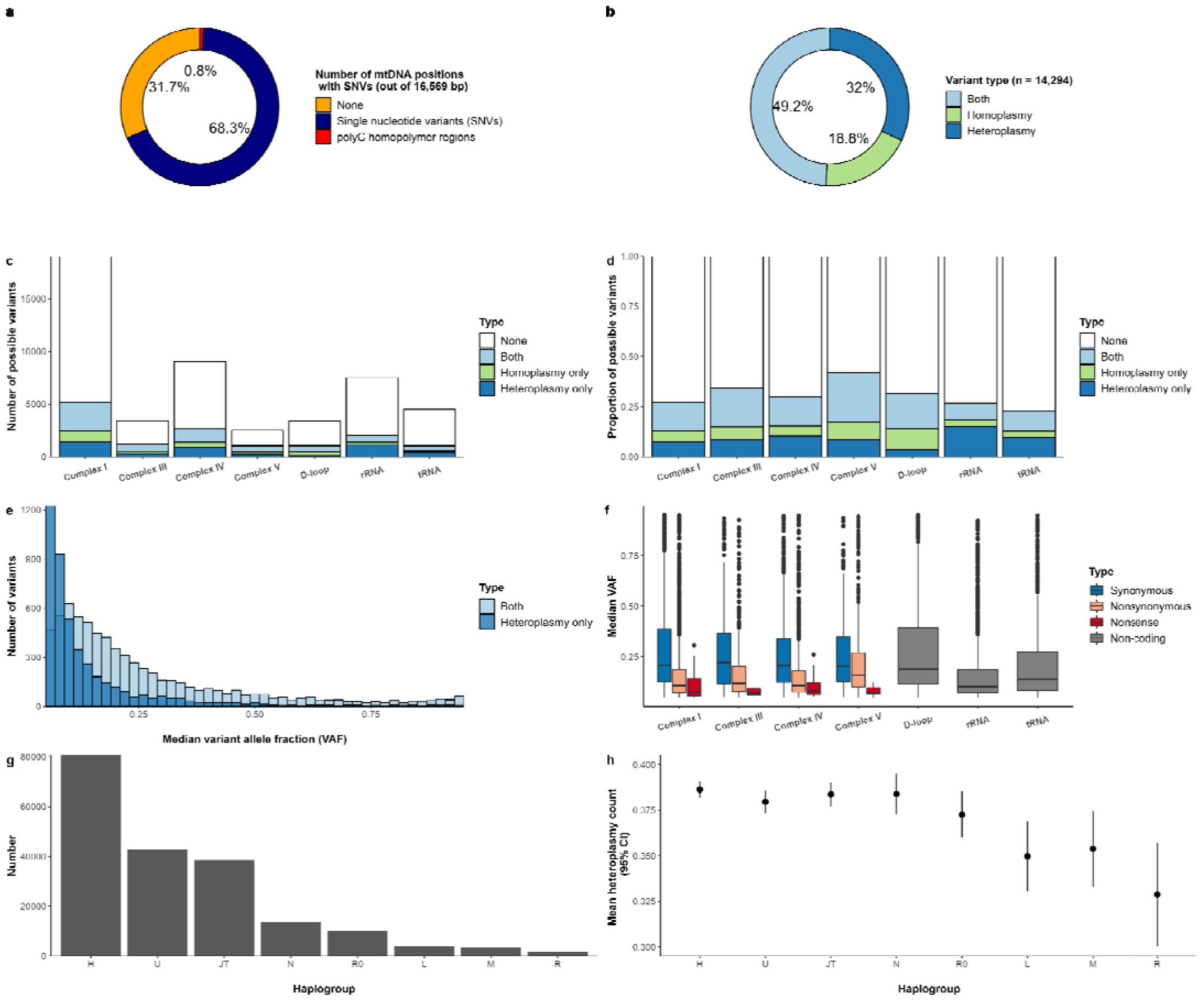
Overview of mitochondrial SNV distributions. **(a)** Proportion of mitochondrial DNA positions with SNVs. **(b)** Proportion of SNVs that are heteroplasmic, homoplasmic, or both. **(c)** Number of possible SNVs that are heteroplasmic, homoplasmic, or both, grouped by protein complex or genic context. **(d)** Bar chart of the proportion of possible SNVs that are heteroplasmic, homoplasmic, or both grouped by protein complex or genic context scaled to 1. **(e)** Histogram of the median variant allele fraction (VAF) for variants seen as only a heteroplasmy or both a hetero- and homoplasmy. The two histograms are overlaid. **(f)** Boxplot of the median VAF by type of mutation. **(g)** Number of participants in each haplogroup. Haplogroups were grouped by phylogenetic similarity into the following: L is L0-L6; M is C, D, E, G, M, Q, Z; N is A, I, N, S, W, X Y; R is B, F, P, R; R0 is R0, HV, V; U is U, K; JT is JT, T; H is H only. **(h)** Mean heteroplasmy count (95% confidence intervals) by haplogroup.

To better understand mitochondrial mutational load, we investigated the characteristics of the heteroplasmic SNVs. We observed a transition-to-transversion (Ti/Tv) ratio of 28.7 for heteroplasmies (**Table 3**) which is also observed in PhyloTree sequences^11^ and likely due to misincorporation by the mitochondrial polymerase gamma^12^. This is in contrast to Ti/Tv ratio in the nuclear genome, which is typically around two^13^. The overall distribution of SNVs by complex/region highlights tRNA genes as less tolerant to variation, with only 23.0% of possible SNVs observed, in contrast to Complex V, with 41.8% of possible SNVs observed (**Fig. 2. c,d**). The overall distribution of the median VAFs for SNVs only seen as heteroplasmies is lower than SNVs seen as both hetero- and homoplasmy (**Fig. 2e**). We hypothesized that SNVs with a functional change would be more likely to have low VAF in a given participant, as these SNVs would likely alter protein function and be under negative selection. Across all participants, we see 4,252 nonsynonymous and 3,950 synonymous SNV sites, with nonsynonymous SNVs having a lower median VAF than synonymous SNVs (**Fig. 2f** and **Table 3**). When stratified by haplogroup (L, M, N, R, R0, U, JT, and H), the number of heteroplasmies significantly differed by haplogroups, with haplogroups L, M, and R, on average, having lower heteroplasmy count compared to haplogroup H **(Fig. 2g, h)**. The variance of heteroplasmy count explained by haplogroups was 0.018%.

**Table 3:** Characteristics of heteroplasmic SNVs by gene type/region.

|  | Length<br>(bp) | Ti/Tv | dN/dS | # Nonsense<br>Mutation Sites<br>(nonsense<br>SNVs / Length<br>*1000) | # Nonsense<br>Mutation<br>Alleles | Mean MLC<br>Score (SE) |
| --- | --- | --- | --- | --- | --- | --- |
| Complex I | 6,349 | 26.8 | 0.79 | 8 (1.3) | 9 | 0.23 (0.0048) |
| Complex III | 1,141 | 25.5 | 1.2 | 6 (5.3) | 11 | 0.16 (0.0066) |
| Complex IV | 3,010 | 24.4 | 1.0 | 28 (9.3) | 57 | 0.29 (0.0070) |
| Complex V | 842 | 19.9 | 1.9 | 7 (8.3) | 8 | 0.06 (0.0040) |
| D-loop | 1,122 | 41.2 | -- | -- | -- | 0.09 (0.0047) |
| tRNA | 1,505 | 26.9 | -- | -- | -- | 0.43 (0.0072) |
| rRNA | 2,513 | 25.2 | -- | -- | -- | 0.55 (0.0057) |
| Unassigned | 87 | 35.1 | -- | -- | -- | 0.28 (0.0300) |
| **Overall** | **16,569** | **28.7** | **1.08** | **49 (4.3)** | **85** | **0.28 (0.0029)** |

Columns are defined as follows: Length is the total number of bases by region or gene type. Protein coding genes are grouped by oxidative phosphorylation (OXPHOS) complexes. The transition-to-transversion ratio (Ti/Tv) is calculated as the number of SNVs with transition changes (A <-> G or T <-> C) divided by the number of SNVs with transversion changes. Nonsynonymous-to-synonymous ratio (dN/dS) is calculated as the number of nonsynonymous mutations divided by synonymous mutations for all protein-coding SNVs. Number of nonsense mutation sites is the sum of all SNVs that cause a stop codon grouped by genes that make up OXPHOS complexes. Number of nonsense mutation alleles is the total number of nonsense mutation alleles present in each OXPHOS complex. Mitochondrial local constraint (MLC) score is calculated from SNVs observed in each gene type or region. The mean and the standard error (SE) of MLC score are reported.

Of the SNVs in Complex I genes, 52% were synonymous SNVs and these genes had the smallest fraction of nonsynonymous SNVs (dN/dS ratio) (**Table 3**). Complex I facilitates the first step of oxidative phosphorylation and deficiencies or inhibition of Complex I lead to increased ROS, reduced NADP levels, and disrupt mitochondrial morphology^14,15^. Interestingly, Complex V genes had the highest dN/dS ratio at 1.9. Complex V, also known as the ATP synthase, is the ATP generating component of the electron transport chain (ETC) and we would expect SNVs in these genes to impact ATP production.

We observed 49 different nonsense heteroplasmic mitochondrial SNVs (encoding stop codons) (**Table 3**) across 85 individuals, with no individual harboring more than one nonsense SNV. One of these variants, m.3308T>G, was also found as a homoplasmic variant at a frequency of ~1:1000, consistent with previous reports of this variant^16^. This variant is likely more tolerated than other nonsense mutations, as variant m.3308T>G is at the start of *MT-ND1*, and the methionine at codon 3 is a likely alternative translation start site, resulting in a loss of only the first 2 amino acids. One additional homoplasmic nonsense variant was found only as a singleton, m.15327C>G near the end of *MT-CYB*. This variant was not reported in gnomAD or Helix database, but is reported in 1 individual in Mitomap, thus demonstrating the strong selection against homoplasmic nonsense SNVs. The majority of the heteroplasmic nonsense mutations occurred at 28 sites in Complex IV genes (*MT-CO1, MT-CO2* and *MT-CO3*), with a marked lower frequency in Complex I genes (**Table 3**). Only 14 nonsense SNVs were seen more than once in the dataset, with the highest frequency SNV seen in 16 individuals. Only 1 individual had VAF > 31%, with a median VAF of 8% compared to 14% for all SNVs, again demonstrating strong selection against these mutations (**Fig. 2f**). There are many well-studied pathogenic mitochondrial SNVs that cause mitochondrial disorders. Of the 91 confirmed pathogenic SNV mutations on Mitomap^16^, 60 unique variants are found in this dataset with 1 variant assigned to two genes, *MT-CO1* and tRNA serine 1 (*MT-TS1*) (**Extended Data Table 1**).

### Association with All-cause Mortality

Mitochondrial heteroplasmic mutations are composed of a combination of inherited and somatic mutations^7^. We would therefore expect that heteroplasmic mutations would increase with age, as has been previously shown^9,17^, as well as with exposure to mutagenic chemicals such as tobacco. Indeed, we observed a significant increase in heteroplasmic SNVs both as a function of age and smoking status (**Fig. 3**). We also observed a significant interaction between age and smoking status (*P* = 1.15 × 10^−5^), likely reflecting the cumulative impact of long-term tobacco use. We did not see a significant association between sex and heteroplasmy count.

**Fig. 3:**
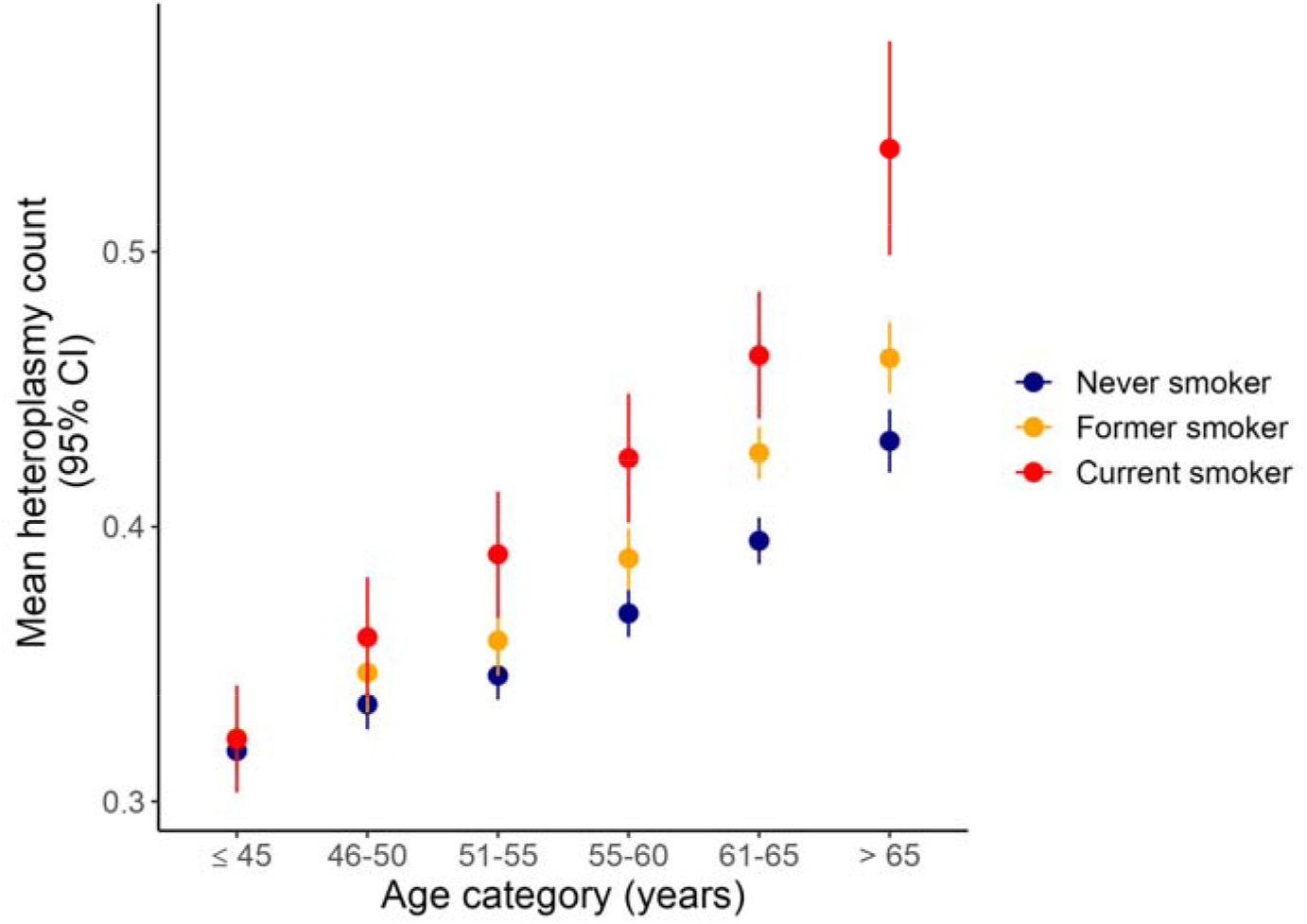
Mean heteroplasmic count by age category and smoking status. Error bars are 95% confidence intervals.

To determine the impact of mitochondrial heteroplasmy on all-cause mortality, we ran Cox proportional hazards models stratified by assessment center and adjusted for age, sex, and smoking status. We observed a 1.50-fold (95% confidence interval [CI] 1.14, 1.98) increased risk of all-cause mortality in individuals with 4 or more heteroplasmies compared to those without any heteroplasmy (**Fig. 4**). This analysis, however, assumes 1) that all heteroplasmies have the same association (effect estimate) with mortality, and 2) it does not directly test the hypothesis that mitochondrial heteroplasmy is causal for increased mortality risk. An alternative hypothesis could be that heteroplasmy count is a non-causal biomarker for some other disease process or environmental exposure, and under this non-causal model, we would predict that the specific functional impact of the heteroplasmic variant would not impact its association with mortality. To distinguish between the causal and non-causal models, we incorporated functional annotations to test whether functional heteroplasmies are driving the association between the number of heteroplasmies and risk of mortality. First, we evaluated whether any of the heteroplasmies with nonsense mutations, which introduce stop codons and therefore, directly decrease protein function, are associated with mortality. The adjusted hazard ratio (aHR) for all-cause mortality was 1.77 (95% CI 1.07, 2.94) and further adjusting for heteroplasmy count marginally attenuated the association (aHR 1.65; 95% CI 0.99, 2.74; **Fig. 4, Extended Data Fig. 3a**), suggesting an association of nonsense mutation and all-cause mortality independent of the number of heteroplasmies, although the confidence interval was wide due to the small number of participants with nonsense mutations (n = 80).

**Fig. 4:**
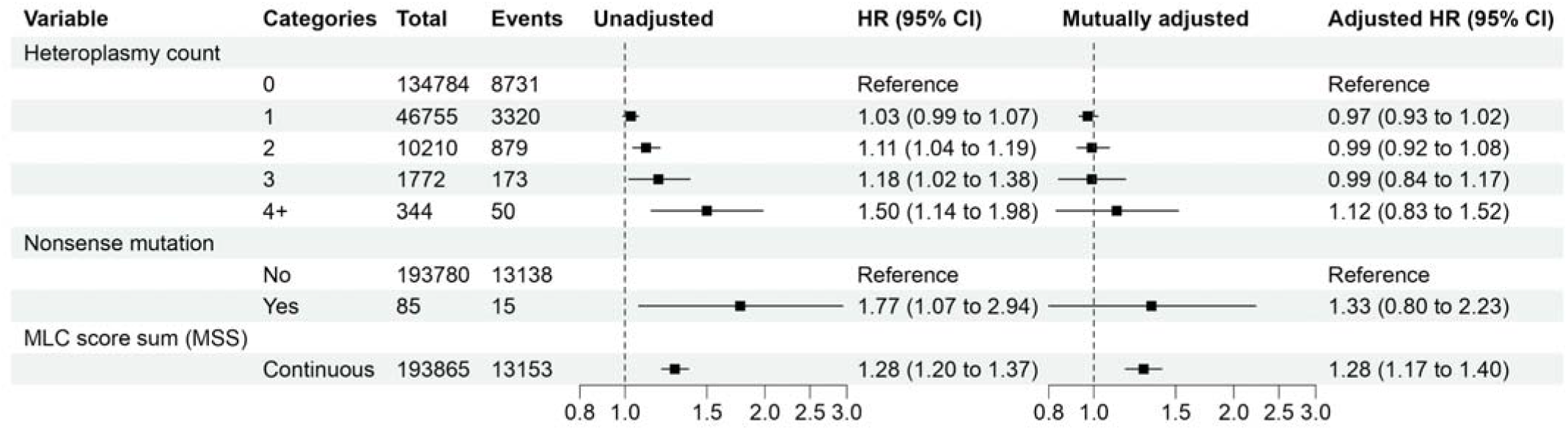
Hazard ratios (95% confidence intervals) for all-cause mortality by heteroplasmy count. Hazard ratios (HR) for the association of all-cause mortality by heteroplasmy count, presence of nonsense mutation, and MLC score sum (MSS) were estimated from separately Cox proportional hazards models (“Unadjusted”) stratified by assessment center and adjusted for age, sex, and smoking status. In addition, heteroplasmy count, nonsense mutation, and MSS were mutually adjusted (“Mutually adjusted”). The numbers of participants and events reflect the number included in the regression analysis.

Second, we incorporated functional annotation into our models to test whether functional heteroplasmies drive the association between the number of heteroplasmies and risk of mortality. We randomly selected one heteroplasmic SNV from each participant, and then tested whether a recently developed annotation metric based on local constraint, Mitochondrial local constraint (MLC) score, had a significant association with all-cause mortality. The MLC score assigns each heteroplasmy a score ranging from 0-1, with 1 being the most constrained SNV, and, therefore, likely to have the most deleterious impact when mutated. MLC score was significantly associated with mortality (adjusted HR comparing an MLC score of 1 to 0 was 1.34; 95% CI 1.21, 1.49, *P* = 5.4 × 10^−8^; **Extended Data Fig. 3**), while continuing to adjust for center (stratification factor), age, sex, smoking status, and number of heteroplasmies. Consistent with the dN/dS ratio, we observed lower MLC scores in Complex I and IV, which appear to be under higher selection constraint (**Table 3**). To capture the impact of multiple heteroplasmies, we generated a MLC score sum (MSS) from all heteroplasmies in each participant. The association between heteroplasmy count and all-cause mortality was no longer significant after adjusting for MSS, while the risk of mortality increased with a higher MSS (aHR for 1-unit increase in MSS 1.28; 95% CI 1.20, 1.37; **Fig. 4**). Furthermore, incorporating the MSS in addition to the presence of nonsense mutations largely explained the effect of heteroplasmy count and nonsense mutation on all-cause mortality (**Fig. 4**). Thus, for all downstream analyses, we used the MSS as a metric that captures the cumulative mutational burden of mitochondrial heteroplasmy. After additionally adjusting for alcohol intake, body mass index (BMI), white blood cell (WBC) counts, and haplogroup, MSS was associated with all-cause mortality in a dose-response manner (aHR 1.28; 95% CI 1.19, 1.37; **Fig. 5**). There ere no significant differences by sex (*P* for interaction = 0.6).

**Fig. 5:**
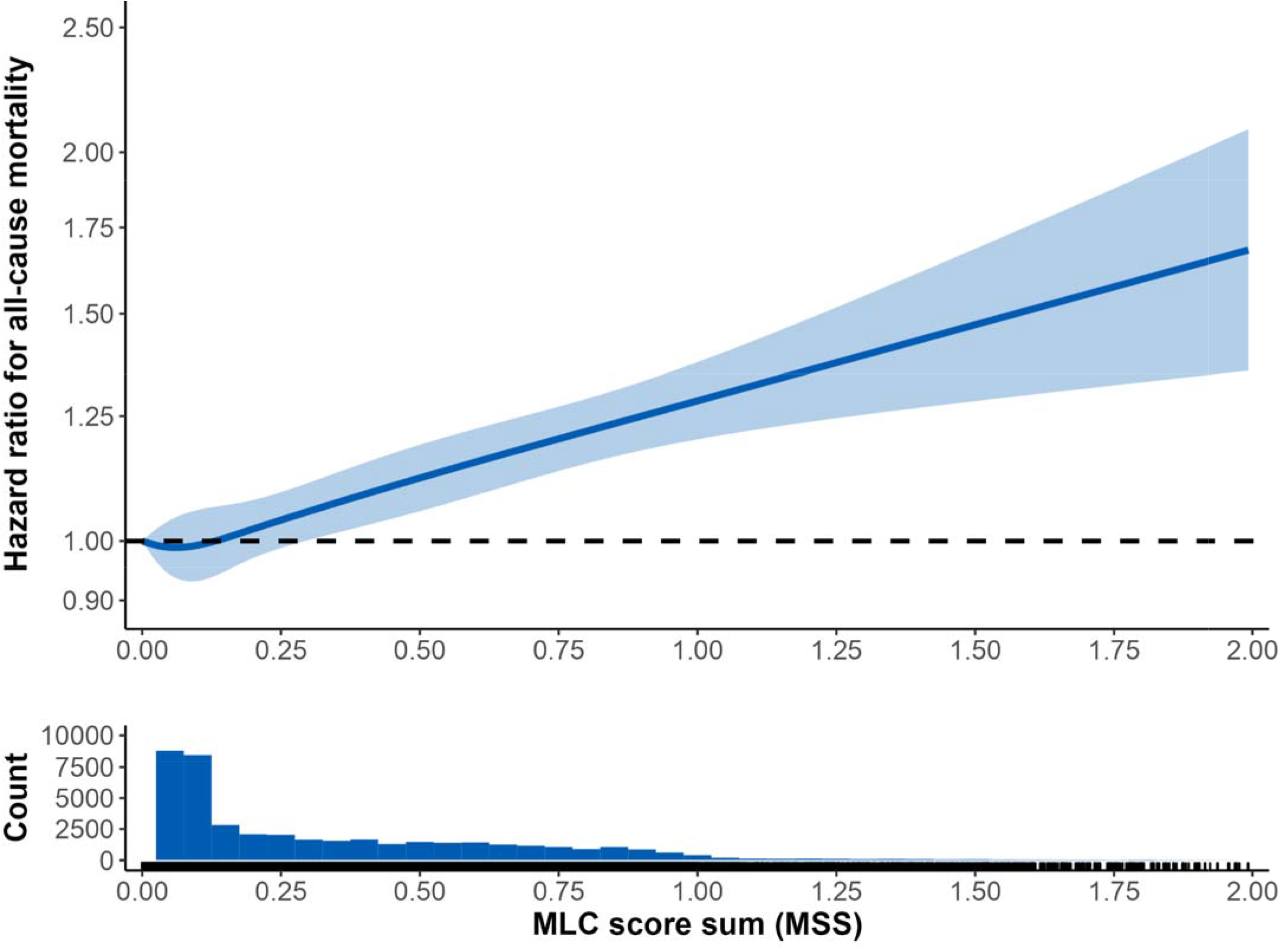
Adjusted dose-response relationship between MSS and all-cause mortality and histogram of MSS among participants with at least 1 heteroplasmy count. Hazard ratios (95% confidence intervals) were derived from Cox proportional hazards models that include MSS as restricted cubic splines with 4 degrees of freedom and were stratified by assessment center and adjusted for age, sex, smoking status, alcohol intake, body mass index, white blood cell count, and haplogroup. 58 participants with MSS greater than 2 were excluded from the plot.

### Association with Cause-specific Mortality

We further investigated if the association with increased mortality was driven by a specific cause of death. When categorizing participants by cause of death using the International Classification of Diseases 10^th^ Revision (ICD-10), MSS was significantly associated with an increased risk of death due to neoplasms (C00– D48), digestive disorders (K20–K93), and external causes (V01–Y89) (**Fig. 6a**). There were no significant differences by sex. Given that the generic category malignant neoplasms (“cancer”) includes many diseases from different organ systems, we further separated solid and hematologic cancers by type (**Fig. 6b**). The MSS was associated with mortality due to lung cancer (aHR 1.56; 95% CI 1.27, 1.91), breast cancer (aHR 1.51; 95% CI 1.06, 2.15), lymphoma (aHR 2.01; 95% CI 1.34, 3.01), and leukemia (aHR 4.97; 95% CI 3.87, 6.39). The associations did not change materially when we restricted the analyses to genetically unrelated participants. Of the external causes of death, the MSS was associated with a higher risk of death due to self-harm (aHR 2.47; 95% CI 1.35, 4.52) but was not associated with death due to accidents (aHR 0.94; 95% CI 0.13, 6.78).

**Fig. 6:**
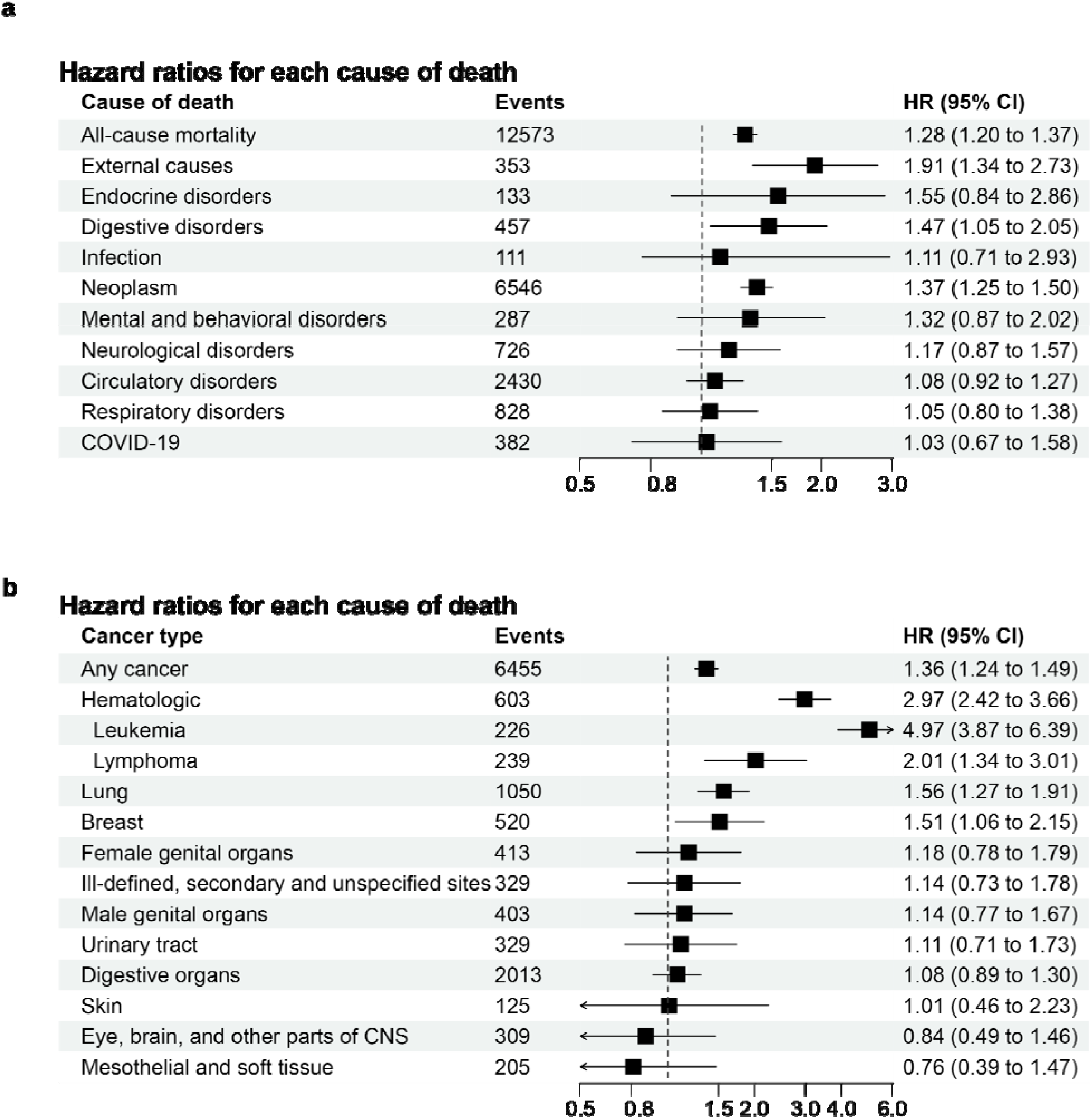
Hazard ratios for each cause of death per 1-unit increase in MLC score sum (MSS). Hazard ratios for (**a**) each specific cause of death, and (**b**) cancer-specific mortality, were estimated using Co proportional hazards models stratified by center and adjusted for age, sex, smoking status, alcohol intake, body mass index, white blood cell count, and haplogroup. Cases of death in fewer than 100 participants (deaths due to benign diseases of the blood [n = 26] or genitourinary disorders [n = 83] in (**a**) and deaths due to cancers of lip, oral cavity, and pharynx [n = 91] or thyroid and other endocrine glands [n = 25] in (**b**)) are not included in the plot. Neoplasm in (**a**) included both benign and malignant neoplasms. Hematologic cancers included cancers of the lymphoid, hematopoietic, and related tissues. Lung cancers included cancers of the respiratory and intrathoracic organs. Abbreviations: CNS, central nervous system. The number of events reflect the number of events in participants included in the regression analysis.

### Cancer Prevalence, Incidence, and Mortality

The association with cancer-specific mortality could arise due to a combination of factors. First, MSS could be a biomarker for cancer presence (prevalence of disease), second, it could be associated with development of cancer (incidence of disease), or third, it could be associated with survival in the presence of disease. We therefore evaluated the associations of MSS with prevalent and incident cancer, as well as survival in those with cancer, for each of the 4 significantly associated cancers (lung, breast, lymphoma, and leukemia) to better distinguish between these potential mechanisms. MSS was associated with both prevalent and incident cases of lung cancer, lymphoma, and leukemia (**Fig. 7a, b**). For breast cancer, the association with prevalent disease was suggestive (adjusted prevalence ratio [aPR] 1.17; 95% CI 0.97, 1.40), whereas there was no association with incident disease (aHR 0.96; 95% CI 0.81, 1.14). The findings suggest a role for MSS as a biomarker of all these cancers and, potentially, the involvement of mitochondrial dysfunction in the development of lung cancer, lymphoma, and leukemia.

**Fig. 7:**
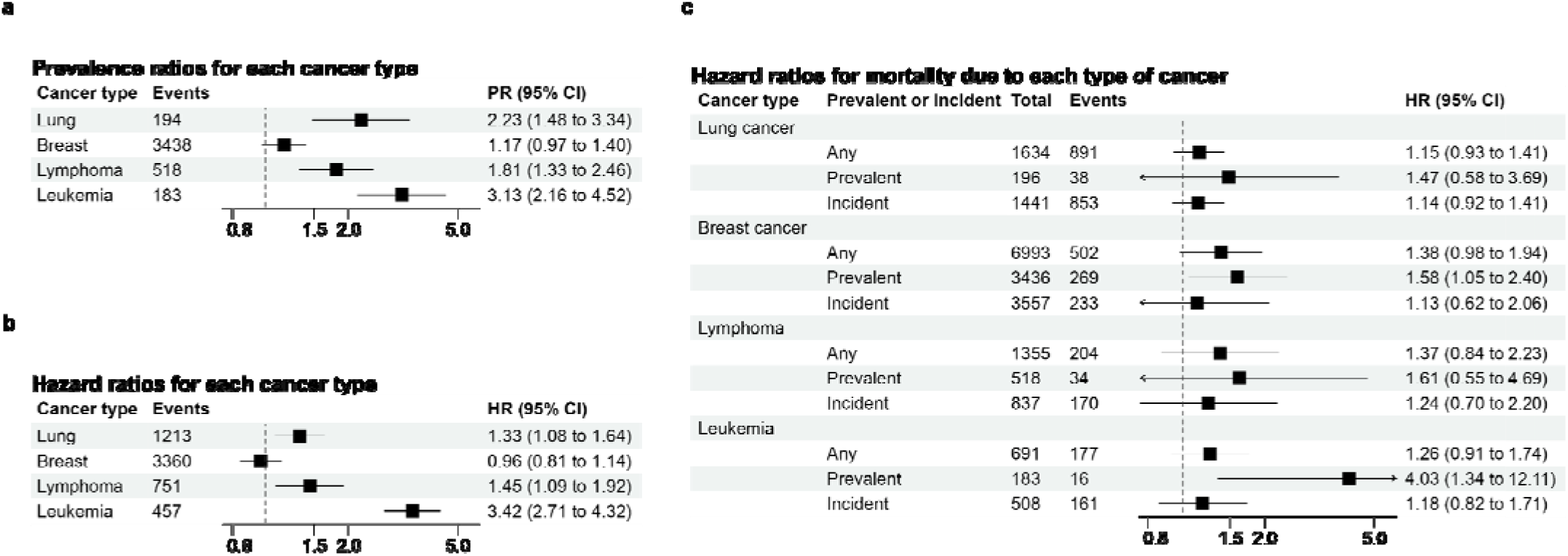
The associations of MSS and cancer prevalence, incidence, and death. (**a**) Prevalence ratios (PRs) and 95% confidence intervals (CIs) for the associations of MSS with prevalent lung cancer, breast cancer, lymphoma, and leukemia were estimated using marginally predicted prevalences from logistic regression models adjusted for age, sex, center, smoking status, alcohol intake, body mass index, white blood cell count, and haplogroup. (**b**) Hazard ratios (HRs) and 95% CI for the associations of MSS with incident lung cancer, breast cancer, lymphoma, and leukemia were estimated using Cox proportional hazards models adjusted as above. For incident cancer, we excluded participants with any history of cancer at the time of blood collection. (**c**) Sub-distribution HRs and 95% CIs for mortality due to lung cancer, breast cancer, lymphoma, and leukemia were estimated using the Fine and Gray method to account for competing events (mortality from other cancers and non-cancer mortality). For cancer events, the analyses were restricted to cancer cases in participants who had prevalent cancer at the time of the UKB visit (“Prevalent”), new cases that were developed during follow up (“Incident”), or had either prevalent or incident cancer (“Any”) of the given type of cancer. The numbers of participants and events reflect the numbers included in the regression analyses.

We further evaluated the risk of mortality from each cancer among those who either had prevalent cancer at the time of UKB visit or had developed cancer (incident cancer) after visit. For all 4 cancers, higher MSS was more strongly associated with mortality among those with prevalent disease, though only significant for leukemia mortality (aHR 4.03; 95% CI 1.34, 12.11) and breast cancer mortality (aHR 1.58; 95% CI 1.05, 2.40) (**Fig. 7c**), suggesting a potential role of MSS as a prognostic marker of these cancers. The associations were similar when we restricted the analyses to genetically unrelated participants and when restricted to participants without extreme values of blood cell counts, which may be indicative of undiagnosed hematological cancers.

### PheWAS and PHESANT Results

Finally, given the varied role that mitochondrial function plays in human health and disease, we examined whether the MSS was associated with a broad array of phenotypes. We first collapsed summary ICD-10 diagnosis codes from UKB inpatient hospital records into 1,618 broad phecodes capturing various disease categories, then tested association between the binary phecodes and MSS in a phenome-wide association study (PheWAS)^18^ (**Extended Data Table 2**). As expected, there was an enrichment for hematological cancers among disease phecodes significantly associated with the MSS, as well as several lung-related phenotypes (lung cancer, bronchitis) (**Fig. 8a**). The MSS was further associated with hematopoietic disease phecodes, such as neutropenia, thrombocytopenia, and aplastic anemia. Notably, PheWAS revealed that the MSS was also associated with sepsis and septicemia phecodes, a finding that was significant even after excluding individuals with leukemia, who often experience sepsis during the course of their disease. A parallel phenome-wide association study was conducted using PHESANT^19^, which additionally tests for associations with a broad array of phenotypes available in the UKB. The MSS was positively associated with several biomarkers of hematopoiesis including immature reticulocytes, platelets, and neutrophil counts, but inversely associated with eosinophil percentage (**Extended Data Table 3**). The MSS was also inversely associated with leukocyte T/S ratios (adjusted, unadjusted, Z-adjusted), markers of relative telomere length. These results are consistent with prior work showing a positive correlation between mtDNA-CN and telomere length^20,21^, further suggesting a role for mitochondrial function in influencing telomere length. Finally, we tested association of the phecodes with identified mtDNA pathogenic variants (**Extended Data Table 1**), using carrier status coded as a synthetic allele to combine the seven selected pathogenic variants. PheWAS of the synthetic allele revealed a statistically significant association to primary/intrinsic cardiomyopathies (phecode 425.1, *P* = 5.0 × 10^−5^; number of cases = 1,014, number of controls = 191,178, number of cases with a pathogenic variant = 10). Notably, m.3243A>G made up a disproportionate frequency among carriers with phecode 425.1 (7/10 cases, 70%) compared to the population frequency (66 / 436 carriers of any pathogenic variant, 15.1%).

**Fig. 8:**
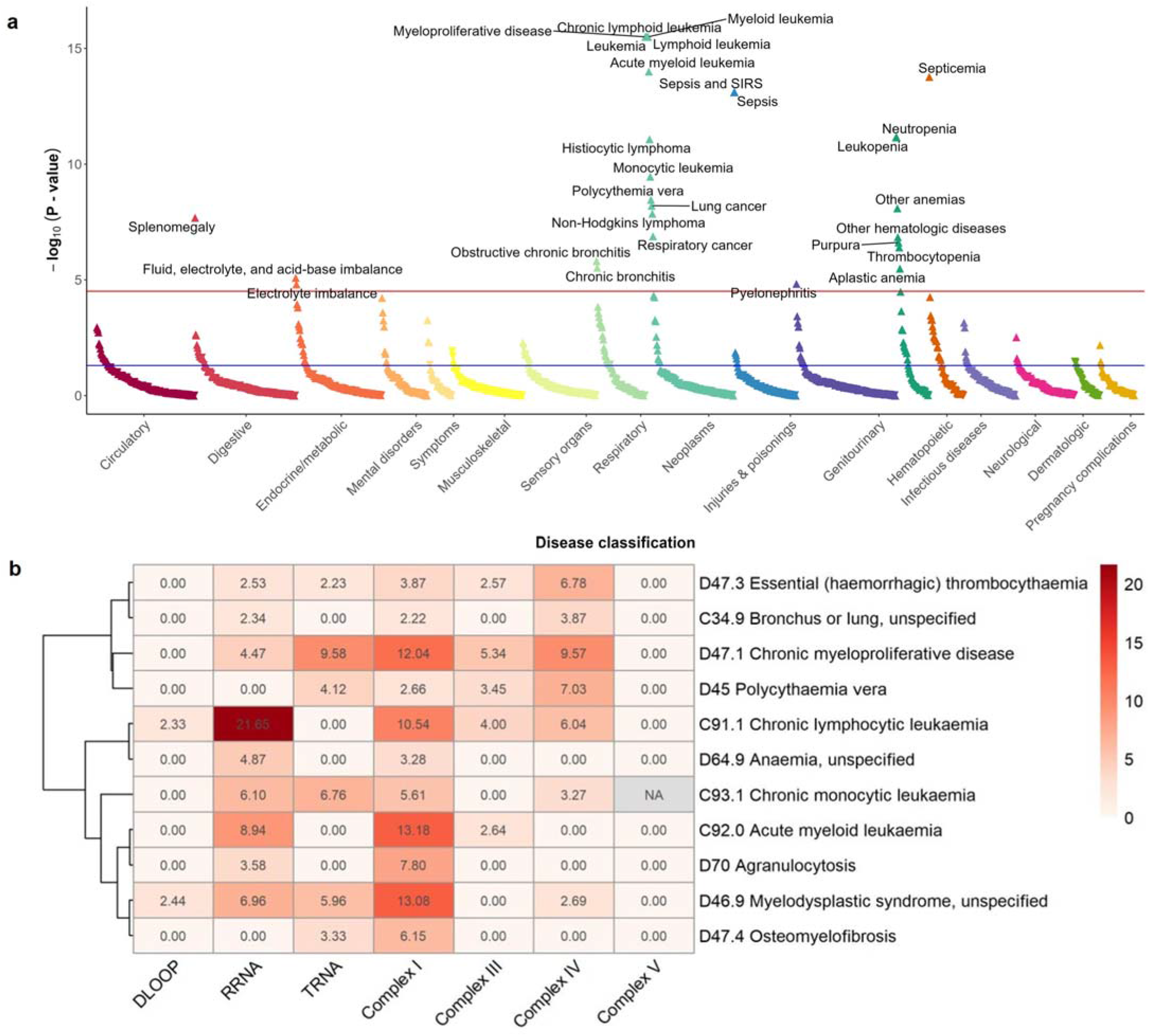
Phenome wide association study (PheWAS) of MSS. **(a)** X-axis indicates disease categories (colors) for phecodes with at least 10 cases, y-axis indicates p-value for association between phecode and MSS. Up/down arrows indicate positive/negative effect direction for association. The blue horizontal line indicates *P* value = 0.05. The red horizontal line indicates *P* value cutoff after Bonferroni correction (3.1 × 10^−5^). Phecodes with *P* values <1.0 × 10^−15^ are plotted at -log_10_(*P* value)) = 15. **(b)** Heatmap displaying the significance (-log_10_(*P* value)) of the association between MSS and ICD 10 codes for cancer (“Chapter II Neoplams”) and hematological diseases (“Chapter III Disease of the blood and blood-forming organs and certain disorders involving the immune mechanism”) stratified by mtDNA region/complex. ICD 10 codes were selected if the significance with the overall MSS was <1×10^−6^, and clustered using Pearson’s correlation. -log_10_(*P* values) >2 (corresponding to *P* values >0.01) were set to 0 for visualization.

Based on the PHESANT results, we selected ICD 10 codes related to cancer and diseases of the blood, and stratified MSS by gene region/complex to see if different mitochondrial functional units were associated with different phenotypes. While high MSSs in Complex I and RRNA were broadly associated across the phenotypes, and the DLOOP and Complex V MSSs were largely not associated with any of the phenotypes, there were also distinct differences observed (**Fig. 8b**). High TRNA MSS was associated with chronic myeloproliferative disease, chronic monocytic leukemia, and myelodysplastic syndrome, and not associated with chronic lymphocytic leukemia (the strongest overall result). Complex III and IV MSSs were most significantly associated with chronic myeloproliferative disease, chronic lymphocytic leukemia, polycythaemia vera, and essential (haemorrhagic) thrombocythaemia (largely Complex IV).

## Discussion

Here we present a detailed examination of the impact of heteroplasmic mitochondrial SNVs, measured in blood, on all-cause mortality and a broad range of phenotypes in almost 200,000 individuals. We identified genes in Complex V as having the most nonsynonymous SNVs and Complex I as having the fewest after adjusting for gene size, along with the lowest dN/dS ratio, as has been observed in a previous large-scale study^6^. Likewise, we see a marked reduction in nonsense SNVs in Complex I genes, consistent with a strong negative selection against mutations that decrease Complex I function relative to the other Complex genes. We identified over 66% of the known pathogenic mutations in our dataset. Some are observed as homoplasmic mutations like 14484T>C, which was predicated to not cause LHON by Bolze et al. based on how frequently it occurred in their dataset (8 in 10,000) and the lack of individuals with the variant having an ICD10 code associated with the disease^22^. We found that the presence of heteroplasmy was associated with all-cause mortality and that a functional score based on local sequence constraint, MSS, was a better predictor variable than the number of heteroplasmies, strongly implicating a causal role for mitochondrial heteroplasmy in all-cause mortality. These results were supported by the observation that, while extremely rare and low VAF, the presence of heteroplasmic nonsense mutations was also associated with increased all-cause mortality risk. In addition to all-cause mortality, MSS was associated with an increased risk of specific causes of death, including neoplasms, digestive disorders, and external causes.

Our most notable finding is the potential of using mitochondrial heteroplasmy as a biomarker of prevalence, incidence, or prognosis of certain cancers. For instance, we found that MSS was associated with the prevalence and incidence of lung cancer, lymphoma, and leukemia. In addition, high MSS was strongly associated with mortality due to leukemia and breast cancer in individuals who had a history of these cancers at the time of blood collection. These results present different models for how mitochondrial dysfunction, measured as the MSS, could increase the risk for cancer mortality. For breast cancer, MSS was not significantly associated with prevalence or incidence of disease, and only appears to impact breast cancer mortality directly. In contrast, the association with lung cancer and lymphoma mortality appears to be largely driven by the association of MSS with both prevalence and incidence of disease, with no significant association with mortality observed among those with disease. Finally, for leukemia, MSS is associated with all three features, with extremely large effect sizes for prevalence, incidence, and mortality due to disease (all PR/HR >3); however, there were relatively few deaths due to leukemia relapse in this dataset (n=17). There have been a limited number of studies linking mitochondrial heteroplasmy and hematological malignancies^23^, with several studies demonstrating increased heteroplasmy due to chemotherapy^24,25^. Intriguingly, while of marginal significance, one study demonstrated that patients with clinically refractive leukemia tended to have higher rates of amino acid altering mutations^24^. These prior studies are consistent with our findings, and suggest that one contribution to higher MSS in individuals with prevalent disease upon DNA collection might be prior chemotherapeutic agent treatment, and further, that successful treatment may be influenced by mitochondrial dysfunction. Accordingly, regular screening for cancer-causing mtDNA mutations or SNVs at highly constrained sequences (high MLC score) could be developed as a prognostic marker for cancer patients and surveillance tool for cancer survivors.

There are a few limitations of our study. First, we only analyzed SNVs at ≥300x depth. Regions with low coverage would appear to have no SNVs in our analysis. This is particularly evident in the 50 bp region downstream of the chrM start site where, even with the circularization approach to mapping in MitoHPC, we still had lower coverage and thus did not count SNVs identified in this region. Second, we only analyzed SNV mutations, as we do not yet have confidence in non-SNV calls (insertions/deletions). Thus, we are missing important sources of mitochondrial DNA variation that are likely to contribute to disease risk. Finally, we have chosen to focus on heteroplasmic variation given the reduced impact of selection on somatic mutation, but fully acknowledge that homoplasmic variation, as well the nuclear genome background on which both heteroplasmic and homoplasmic variants fall, may also have important impacts on disease risk.

The first disease causing SNV of mtDNA was identified in 1988^26,27^. Now with next-generation sequencing scientists can query the full mitochondrial genomes of hundreds of thousands of individuals to investigate how mtDNA mutations contribute to common diseases. We found that mitochondrial heteroplasmic mutational burden, measured as the sum of a sequence-based local constraint score, is associated with all-cause and cancer-specific mortality. Moreover, this score has the potential to be used as a prognostic marker, identifying those at risk of both developing disease, as well as those who may benefit from more careful monitoring or intervention post disease onset.

## Data Availability

All data in the present study were obtained directly from the UK Biobank.

## Methods

### UK Biobank WGS data

The UK Biobank is a large population-based prospective study of 500,000 participants aged between 40 to 69 years recruited across the United Kingdom from 2006 to 2010^10^. The UK Biobank collects extensive phenotypic and genotypic data on participants, which are used in our analysis. We included in our analysis 199,909 participants who underwent WGS of the DNA from the blood draw and consented to be in the study.

### MitoHPC on DNA Nexus (heteroplasmy and CN)

All UK Biobank data was processed on the DNA Nexus server. MitoHPC uses Mutect2^28,29^ for variant identification. WGS CRAM files were used for variant calling, haplogroup identification, and contamination checks. We implemented a heteroplasmy allele frequency of 5%, meaning that variant alleles at a frequency 5-95% in an individual are counted as heteroplasmic. Alleles less than 5% or greater than 95% are counted as homoplasmic. We used SAMtools^30^ to generate read count and coverage information for mtDNA-CN calculations, using the command ‘samtools idxstats’. Documentation on how to run MitoHPC on DNA Nexus server is available here: https://github.com/dpuiu/MitoHPC/blob/main/docs/DNAnexus_CLOUD.mdOm

### Mitochondrial local constraint (MLC) score

The mitochondrial local constraint (MLC) score is a measure of local tolerance to base or amino acid substitutions. It was calculated for every possible mtDNA single nucleotide variant (SNV) by applying a sliding window method (Lake et al, manuscript in preparation). In brief, starting from position m.1 a window of length k is drawn and the observed:expected (oe) ratio of substitutions in gnomAD within the window and its 90% confidence interval (CI) is calculated. The window start position is then moved by 1 bp, and this process is repeated until a start position of m.16569 is reached. For positions in protein genes only missense variants are included in calculations to restrict to amino acid substitutions, while for all other positions all base substitutions are used. The mean oe ratio 90% CI upper bound fraction (OEUF) of all k windows overlapping each position is computed, and percentile ranked to achieve a score range of 0-1 where 1 is most constrained position and 0 is least constrained. A MLC score is obtained for every mtDNA SNV as follows: non-coding, RNA and missense variants are assigned their positional score, and non-missense in protein genes are assigned scores based on the OEUF value of the variant class with synonymous, stop gain, and start/stop lost being assigned scores of 0.0, 1.0, and 0.70 respectively. Variants with higher scores are predicted to be more deleterious.

### Sample QC and Variant Filtering

We ran 200,000 WGS samples in the UK Biobank (UKB) database through MitoHPC. Of those 199,919 samples had outputs from MitoHPC variant calling and of those we calculated mtDNA copy number (mtDNA-CN) in 199,910 samples (**Extended Data Fig. 1**). The median nuclear genomic coverage for WGS samples in the UKB is 35x and the median mtDNA coverage is 1113x. This allowed us to confidently call heteroplasmy with variant allele fractions (VAF) as low as 5%.

MitoHPC outputs various metrics for assessing sample quality, allowing us to remove low quality samples prior to analysis. MitoHPC uses Mutect2 for variant calling and outputs variant annotations that we used to filter for high quality variants. We excluded variants with read depth < 300 and those flagged as base quality, strandedness, slippage, weak evidence, germline, position flags in the FILTER column of the VCF. We further excluded heteroplasmic variants at poly-C homopolymer regions on the mitochondrial chromosome and excluded INDELs. INDELs are often found at homopolymer regions and due to the nature of these regions, are challenging to accurately call heteroplasmies^9^. Nuclear-encoded mitochondrial sequences (NUMTs) can contribute to false positive mtDNA heteroplasmy calls. However, the nuclear genome coverage is ~31x lower than the mitochondrial genome coverage and by implementing at mtDNA-CN cutoff, we reduce the influence of NUMTs on our data. We excluded samples based on a few criteria: potential mitochondrial contamination, 2 or more variants belonging to a different mitochondrial haplogroup, multiple variants predicted to be NUMTs, low minimum base coverage, and low mean base coverage (**Extended Data Fig. 2**) which resulted in 2501 participants being excluded. Since mitochondrial heteroplasmy has previously been shown to be affected by low mtDNA-CN^6^, we removed participants with mtDNA-CN less than 40 (n = 3580; **Extended Data Fig. 1** and **Extended Data Fig. 2**,**c**). Some samples met multiple exclusion criteria. We found that 358 participants had a heteroplasmic count above 5, with 175 of them identified as contaminated. We removed the remaining high heteroplasmic samples as these appeared to be outliers in our dataset with potentially unidentified contamination. Finally, we were left with 194,871 participants to use for downstream analysis. We plotted the distribution of mtDNA-CN and heteroplasmic count for each participant, to visualize how these cutoffs affect the data (**Extended Data Fig. 2**).

### Statistical Analysis

The UK Biobank is linked to national death registries and provides information on date of death and cause of death. Primary cause of death was coded using the ICD-10 codes and classified into 12 categories (infection [A00–B00, L00–L08]; neoplasm [C00–D48]; benign disease of the blood [D50–D89]; endocrine disorders [E00–E90]; mental and behavioral disorders [F00–F89]; neurological disorders [G00–G99]; circulatory disorders [I05–I89]; respiratory disorders [J09–J99]; digestive disorders [K20–K93]; genitourinary disorders [N00–N98]; COVID-10 [U07]; and external causes [V01–Y89], **Extended Data Table 4**). Neoplasm-related mortality was further separated into benign neoplasm [D00–D48] and malignant neoplasm [C00–C97], which was further categorized by type of cancer.

The UK Biobank also provides information on cancer diagnosis by linkage to national cancer registries, including type of cancer, date of cancer diagnosis, and age at cancer diagnosis. The type of cancer is coded using ICD-9 or ICD-10 codes, which we used to categorize into 15 types of cancer by organ system (**Extended Data Table 5**). For malignant neoplasms, stated or presumed to be primary, of lymphoid, hematopoietic and related tissue (“hematologic cancers”, ICD-10 C81–C96, ICD-9 200–208), we further identified individuals with lymphoma (ICD-10 C81–C86, ICD-9 200–202) or leukemia (ICD-10 C91–C95, ICD-9 204–208). ICD-10 codes from the death registry were also used to identify participants who were diagnosed with cancer. Skin cancer only included melanoma cases and breast cancer cases were restricted to women. We used date of cancer diagnosis (from national cancer registries) or date of death (from the death registry), whichever came first, as the date of cancer diagnosis in the analysis. Cancers diagnosed before the UKB visit were defined as prevalent cancers and cancers diagnosed after the UKB visit were defined as incident cancers.

Survival analyses for all-cause mortality and disease-specific mortality were performed using Cox proportional hazards model to estimate the hazard ratios (HRs) and their corresponding 95% CIs. Each participant was followed from the date of visit to the date of death or to November 12, 2021 (administrative censoring), whichever came first. For COVID-19, on the other hand, we allowed late entries where follow-up started from the first date of COVID-19 mortality on record. Models for evaluating the association between heteroplasmy count and all-cause mortality were stratified by assessment center and were adjusted for age using restricted cubic splines with 4 degrees of freedom, sex, and smoking status (never, former, or current). Models evaluating the associations of MLC score with all-cause mortality and disease-specific mortality were further adjusted for alcohol intake (never, former, or current), BMI (continuous), WBC counts (continuous), and haplogroup (L, M, N, R, R0, U, JT, and H). To evaluate the non-linear association between MLC score and all-cause mortality, MLC score was modeled using restricted cubic splines with 4 degrees of freedom.

In addition, we evaluated the risk of mortality from cancers of the lung, breast, lymphoma, and leukemia by MLC score using a proportional sub-distribution hazards model^31^ to account for competing events (mortality from other cancers and non-cancer deaths, separately). Individuals who developed cancer during the study period (incident cancer) were followed from the date of cancer diagnosis whereas individuals with prevalent cancer at the time of assessment visit were followed from the date of visit (late entry) to account for immortal person time. The competing risk models were adjusted for age (restricted cubic splines with 4 degrees of freedom), sex, assessment center, smoking status, alcohol intake, BMI, WBC counts, and haplogroup.

We further estimated the prevalence ratios (PRs) of the 4 types of cancers using logistic regression models. We used marginally predicted probabilities to calculate PRs associated with a 1-unit increase in MLC score adjusted for age (restricted cubic splines with 4 degrees of freedom), sex, assessment center, smoking status, alcohol intake, BMI, WBC counts, and haplogroup. The corresponding 95% CIs were estimated using the delta method. For the same 4 types of cancers, HRs and their 95% CIs were estimated using Cox proportional hazards models stratified by assessment center and adjusted for the same covariates. Participants with any cancer diagnosis prior to date of visit were excluded from the analyses of incident cancers. Participants were followed from the date of visit to date of cancer diagnosis, date of death, or administrative censoring, whichever came first.

As the UKB includes genetically related participants, we additionally repeated the analyses restricting to genetically unrelated participants (as defined in the UK Biobank as variable used.in.pca.calculation). Moreover, we further restricted the study population to participants without extreme values of red blood cell (RBC), WBC, platelets, and differential WBC counts. Extreme values of total and differential WBC counts were based on visual inspection and defined as log(RBC + 1) ≤1.4, log(RBC + 1) ≥2, log(WBC + 1) ≤1.25, log (WBC + 1) ≥3, platelets ≤ 10,000/µL, platelets ≥ 500,000/µL, log(neutrophils + 1) ≤0.75, log(neutrophils + 1) ≥2.75, log(lymphocytes + 1) ≤0.10, log(lymphocytes + 1) ≥2, log(monocytes + 1) ≥0.9, log(eosinophils + 1) ≥0.75, or log(basophils + 1) ≥0.45.

### PheWAS

We used the R package PheWAS^18^ to collapse summary diagnoses indicated by primary and secondary ICD-10 disease codes from hospital inpatient records into distinct phecodes comprised of cases and controls for a variety of disease categories. The analysis was restricted to all participants with a MSS including 193,866 individuals and 1,618 phecodes with at least one case. This number was used for correction for multiple testing using the Boneferroni method. A logistic regression was run for each phecode regressing the binary phecode onto MSS residuals from a linear regression model adjusting for age, sex, and smoking status. Additional covariates in the logistic regression include age incorporated as a natural spline with four degrees of freedom, sex, and assessment center. Age was modeled using restricted splines with four degrees of freedom. Firth correction was used to account for unbalanced case and controls when the logistic regression p-value was ≤0.0005 using the R package logistf^32^.

## PHESANT

We used the PHEnome Scan ANalysis Tool (PHESANT)^57^ to identify MSS associated phenotypes in the UKB. Briefly, we tested for the association of mtDNA-CN with ~30,000 traits (**Extended Data Table 3**), using MSS residuals from a linear regression model adjusting for age, sex, and smoking status, with age incorporated as a natural spline with four degrees of freedom. PHESANT was then run with the residuals with additional adjustment for age, sex, and assessment center.

## Data and code availability

Data is available through application to the UK Biobank. Code is available on our github repository: https://github.com/ArkingLab.

## Extended Data

**Extended Data Table 1.** 60 unique pathogenic variants.

**Extended Data Table 2.** A phenome-wide association study (PheWAS) results for association with MSS.

**Extended Data Table 3.** PHESANT results for association with MSS.

**Extended Data Table 4.** ICD-10 codes for each cause of death.

**Extended Data Table 5.** ICD-10 and ICD-9 codes for each type of cancer.

**Extended Data Fig. 1.** Flowchart.

**Extended Data Fig. 2.** Data QC and participant exclusion criteria.

**Extended Data Fig. 3.** Hazard ratios (95% confidence intervals) for all-cause mortality by heteroplasmy count adjusted for nonsense mutation and MLC score.

## Acknowledgements

This research was conducted using the UK Biobank Resource under Application Number 17731. This work was supported by National Heart, Lung and Blood Institute, National Institutes of Health (NIH) grant R01HL144569, National Health and Medical Research Council Fellowship (1159456) and American Australian Association Scholarship. The content is solely the responsibility of the authors and does not necessarily represent the official views of the NIH.

## Author contributions

S.L.B., Y.S.H, W.S, D.P., V.P., D.E.A. performed the analyses; N.J.L. and M.L. developed the MLC score; N.P. and E.G. aided interpretation of the study data; S.L.B., Y.S.H., V.P., and D.E.A. drafted the manuscript; all authors read and approved the final manuscript. E.G. and D.E.A supervised and managed the study. S.L.B. and Y.S.H contributed equally to this work.

## Additional Information

Supplementary Information is available for this paper.

Correspondence and requests for materials should be addressed to Dan E. Arking.

Dan E. Arking

McKusick-Nathans Institute

Department of Genetic Medicine

Johns Hopkins University School of Medicine

733 N. Broadway

Miller Research Building, Room 459

Baltimore, MD 21205

Table presented in text for initial submission.

## Figure Legends

Figures and Figure legends are presented in the text for initial submission.

