## Supplementary figures and images for "Deleterious heteroplasmic mitochondrial mutations increase risk of overall and cancer-specific mortality"

### Extended Data Fig. 1

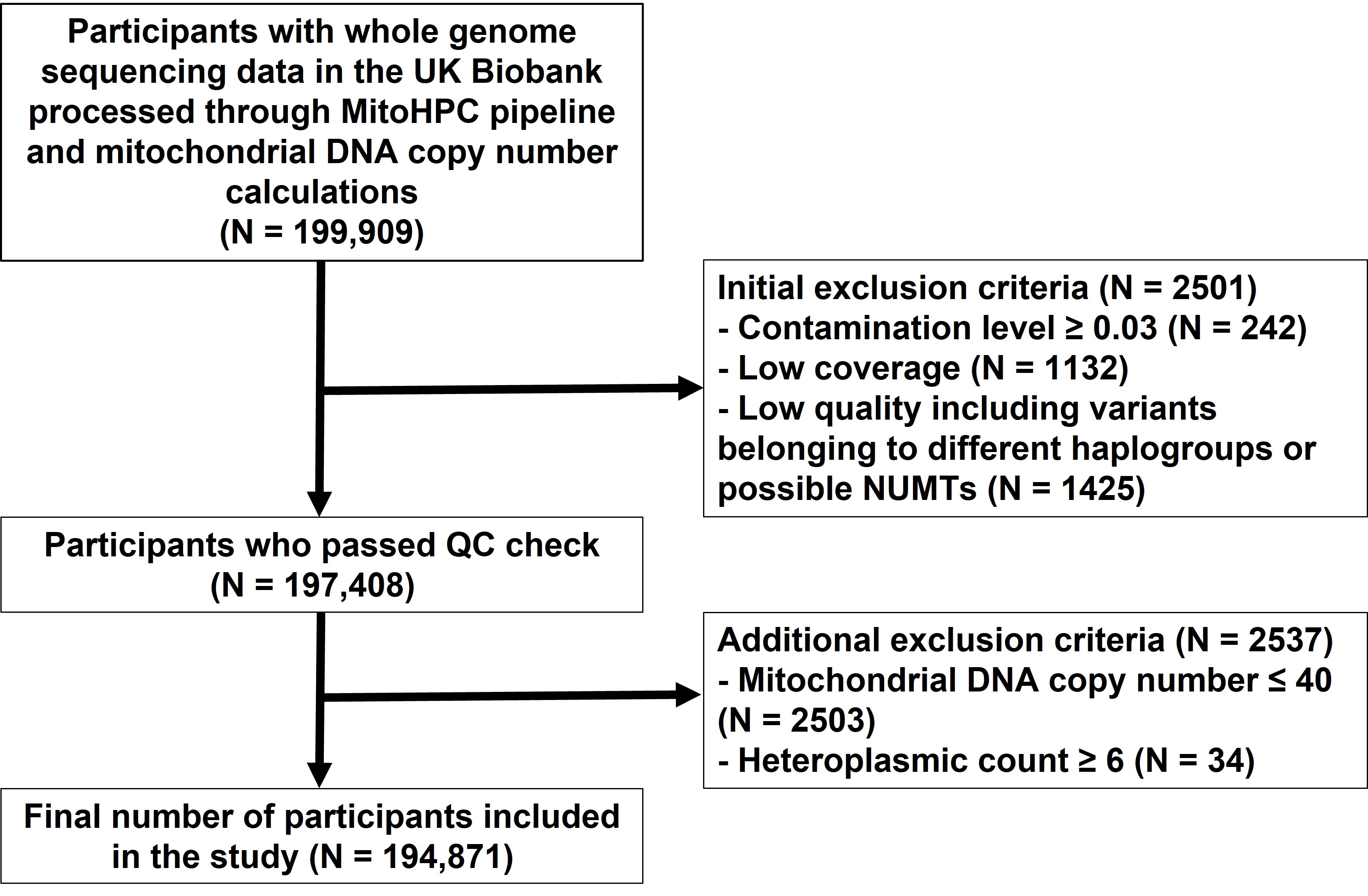

### Extended Data Fig. 2

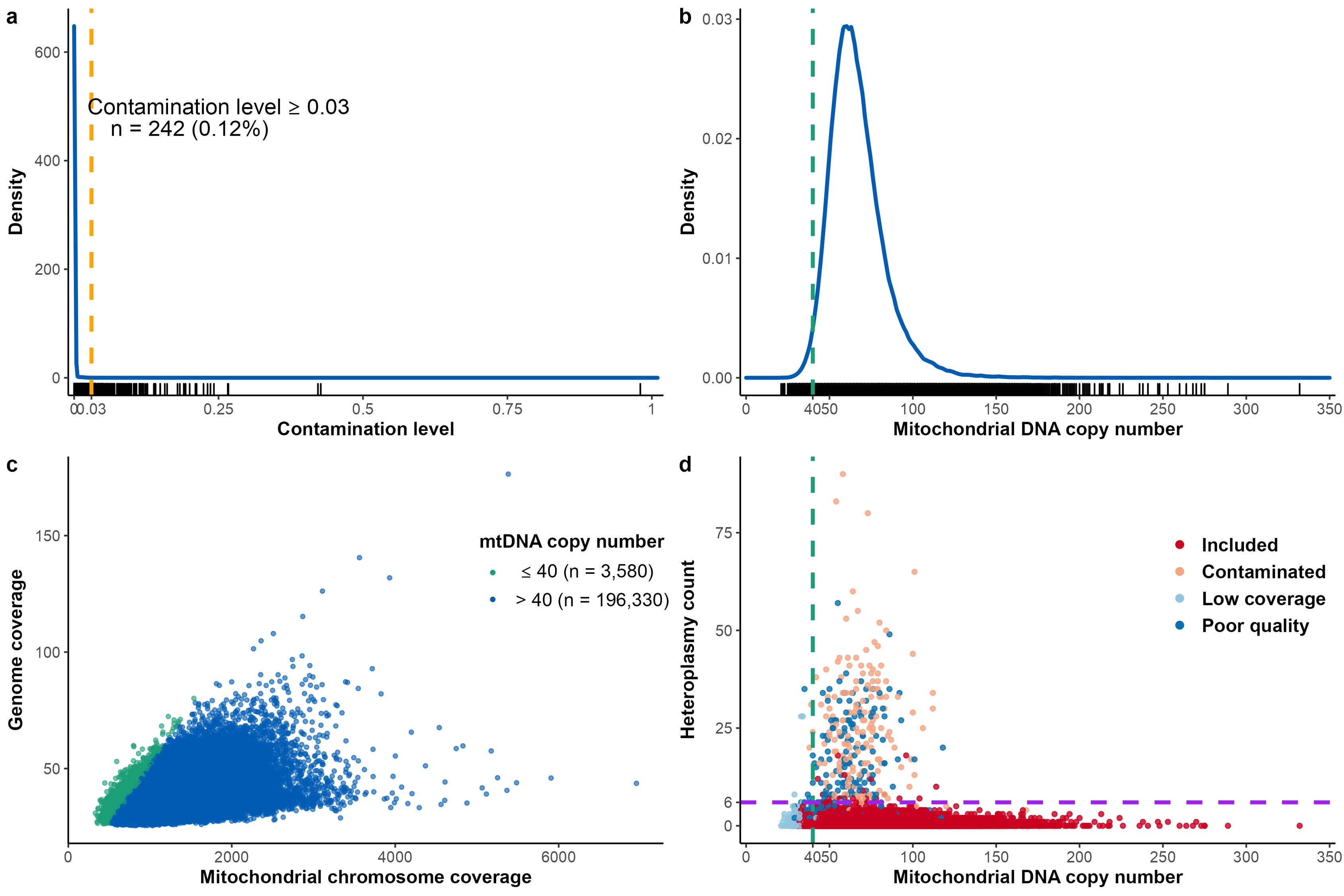

### Extended Data Fig. 3

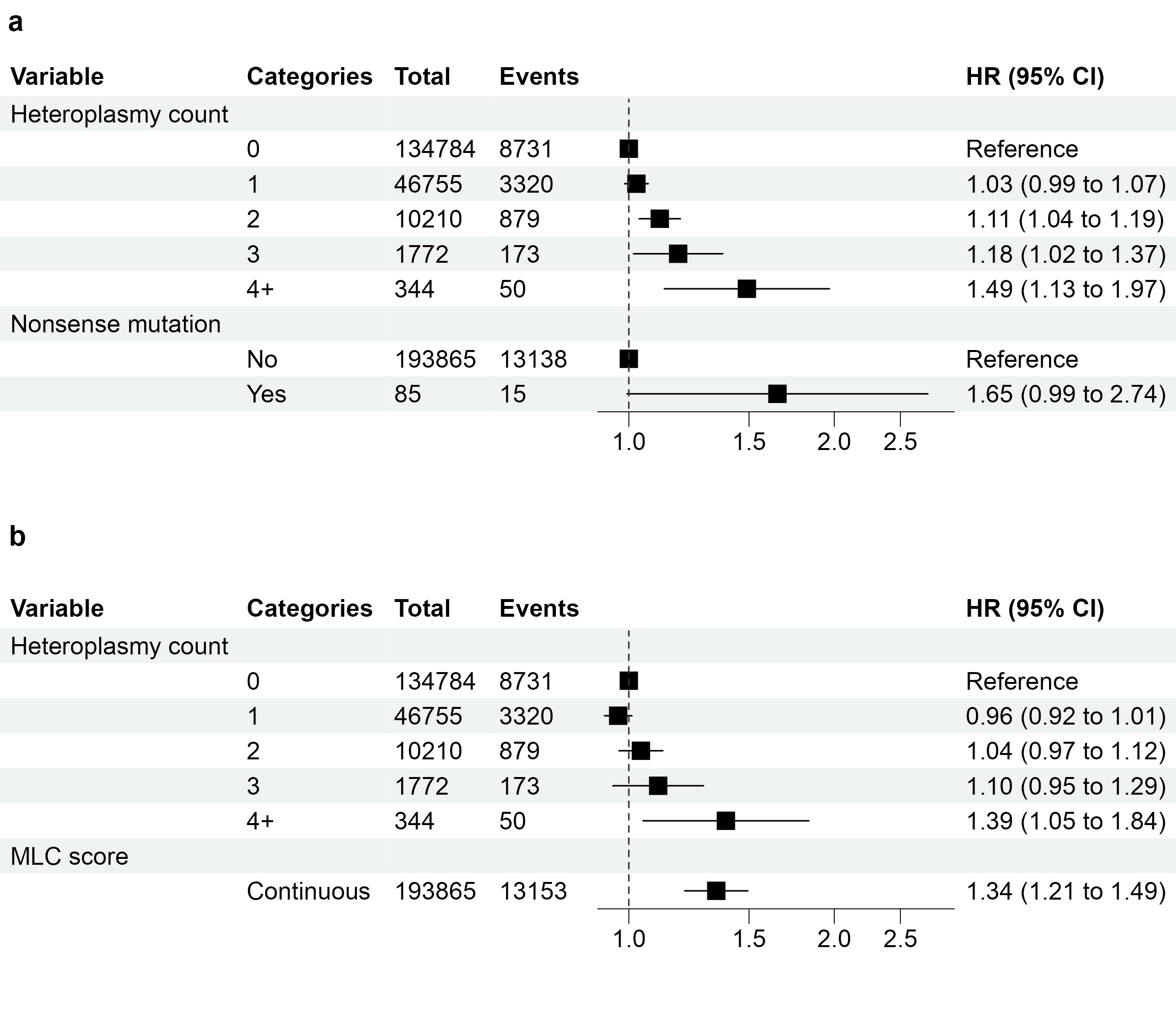
