## Extended Data for "Deleterious heteroplasmic mitochondrial mutations increase risk of overall and cancer-specific mortality"

**List of Extended Data Tables**

**Extended Data Table 1.** **60 unique pathogenic variants.**

| **Position_Ref_Alt** | **GENE** | **COMPLEX/REGION** | **MLC score** | **Median Heteroplasmy** **VAF** | **MitoMap-Assigned Gene** | **MitoMap-associated Diseases** |
| --- | --- | --- | --- | --- | --- | --- |
| 10010_T_C | TRNG | TRNA | 0.46725813 | 0.099 | MT-TG | PEM |
| 10158_T_C | ND3 | I | 0.519283 | 0.0785 | MT-ND3 | Leigh Disease / MELAS |
| 10191_T_C | ND3 | I | 0.55389583 | 0.064 | MT-ND3 | Leigh Disease / Leigh-like Disease / ESOC |
| 10197_G_A | ND3 | I | 0.60869696 | 0.095 | MT-ND3 | Leigh Disease / Dystonia / Stroke / LDYT |
| 10663_T_C | ND4L | I | 0.87192951 | 0.124 | MT-ND4L | LHON |
| 11778_G_A | ND4 | I | 0.8857505 | 0.307 | MT-ND4 | LHON / Progressive Dystonia |
| 12201_T_C | TRNH | TRNA | 0.33909711 | 0.201 | MT-TH | Maternally inherited non-syndromic deafness |
| 12276_G_A | TRNL2 | TRNA | 0.4613133 | 0.052 | MT-TL2 | CPEO |
| 12315_G_A | TRNL2 | TRNA | 0.42633834 | 0.068 | MT-TL2 | CPEO / KSS / possible carotid atherosclerosis risk, trend toward myocardial infarction risk |
| 13042_G_A | ND5 | I | 0.92953709 | 0.292 | MT-ND5 | Optic neuropathy/ retinopathy/ LD |
| 13051_G_A | ND5 | I | 0.87739151 | 0.079 | MT-ND5 | LHON |
| 13379_A_G | ND5 | I | 0.6536001 | 0.076 | MT-ND5 | LHON |
| 13513_G_A | ND5 | I | 0.63292896 | 0.055 | MT-ND5 | Leigh Disease / MELAS / LHON-MELAS Overlap Syndrome / negative association w Carotid Atherosclerosis |
| 14459_G_A | ND6 | I | 0.83463094 | 0.053 | MT-ND6 | LDYT / Leigh Disease / dystonia / carotid atherosclerosis risk |
| 14484_T_C | ND6 | I | 0.43828837 | 0.177 | MT-ND6 | LHON |
| 14487_T_C | ND6 | I | 0.42111775 | 0.079 | MT-ND6 | Dystonia / Leigh Disease / ataxia / ptosis / epilepsy |
| 14568_C_T | ND6 | I | 0.12300078 | 0.227 | MT-ND6 | LHON |
| 14674_T_C | TRNE | TRNA | 0.39945078 | 0.122 | MT-TE | Reversible COX deficiency myopathy |
| 14709_T_C | TRNE | TRNA | 0.75291206 | 0.07 | MT-TE | MM+DMDF / Encephalomyopathy / Dementia+diabetes+ophthalmoplegia |
| 1494_C_T | RNR1 | RRNA | 0.8143521 | 0.628 | MT-RNR1 | DEAF |
| 1555_A_G | RNR1 | RRNA | 0.82189631 | 0.1985 | MT-RNR1 | DEAF; autism spectrum intellectual disability; possibly antiatherosclerotic |
| 1630_A_G | TRNV | TRNA | 0.35152997 | 0.359 | MT-TV | MNGIE-like disease / MELAS |
| 1644_G_A | TRNV | TRNA | 0.31905969 | 0.1665 | MT-TV | Leigh Syndrome / HCM / MELAS |
| 3243_A_G | TRNL1 | TRNA | 0.85509083 | 0.089 | MT-TL1 | MELAS / Leigh Syndrome / DMDF / MIDD / SNHL / CPEO / MM / FSGS / ASD / Cardiac+multi-organ dysfunction |
| 3243_A_T | TRNL1 | TRNA | 0.85509083 | 0.075 | MT-TL1 | MM / MELAS / SNHL / CPEO |
| 3256_C_T | TRNL1 | TRNA | 0.76356449 | 0.099 | MT-TL1 | MELAS; possible atherosclerosis risk |
| 3271_T_C | TRNL1 | TRNA | 0.71344076 | 0.074 | MT-TL1 | MELAS / DM |
| 3302_A_G | TRNL1 | TRNA | 0.49269721 | 0.073 | MT-TL1 | MM |
| 3303_C_T | TRNL1 | TRNA | 0.46490434 | 0.096 | MT-TL1 | MMC |
| 3376_G_A | ND1 | I | 0.29947492 | 0.057 | MT-ND1 | LHON MELAS overlap |
| 3460_G_A | ND1 | I | 0.88481502 | 0.107 | MT-ND1 | LHON |
| 3635_G_A | ND1 | I | 0.60815378 | 0.1065 | MT-ND1 | LHON |
| 3697_G_A | ND1 | I | 0.90949967 | 0.117 | MT-ND1 | MELAS / Leigh Syndrome / LDYT / BSN |
| 3700_G_A | ND1 | I | 0.88840606 | 0.1095 | MT-ND1 | LHON |
| 3733_G_A | ND1 | I | 0.37911159 | 0.075 | MT-ND1 | LHON |
| 3890_G_A | ND1 | I | 0.93288672 | 0.059 | MT-ND1 | Progressive Encephalomyopathy / Leigh Syndrome / Optic Atrophy |
| 4300_A_G | TRNI | TRNA | 0.7288913 | 0.3265 | MT-TI | MICM |
| 4332_G_A | TRNQ | TRNA | 0.49864204 | 0.053 | MT-TQ | Encephalopathy / MELAS |
| 4450_G_A | TRNM | TRNA | 0.67777174 | 0.147 | MT-TM | Myopathy / MELAS / Leigh Syndrome |
| 5521_G_A | TRNW | TRNA | 0.43683988 | 0.06 | MT-TW | Mitochondrial myopathy |
| 5650_G_A | TRNA | TRNA | 0.57580421 | 0.073 | MT-TA | Myopathy |
| 5690_A_G | TRNN | TRNA | 0.81486511 | 0.095 | MT-TN | CPEO+ptosis+proximal myopathy |
| 5703_G_A | TRNN | TRNA | 0.7905124 | 0.101 | MT-TN | CPEO / MM |
| 5728_T_C | TRNN | TRNA | 0.77219506 | 0.06 | MT-TN | Multiorgan failure / myopathy |
| 616_T_C | TRNF | TRNA | 0.49894381 | 0.161 | MT-TF | Maternally inherited epilepsy / mito tubulointerstitial kidney disease (MITKD) / Gitelman-like syndrome |
| 7445_A_G | COX1 | IV | 0.55338282 | 0.165 | MT-TS1 precursor | SNHL |
| 7445_A_G | COX1 | IV | 0.55338282 | 0.165 | MT-CO1 | SNHL |
| 7497_G_A | TRNS1 | TRNA | 0.40126139 | 0.084 | MT-TS1 | MM / EXIT |
| 7510_T_C | TRNS1 | TRNA | 0.52275333 | 0.585 | MT-TS1 | SNHL |
| 7511_T_C | TRNS1 | TRNA | 0.52731004 | 0.166 | MT-TS1 | SNHL/Deafness |
| 8344_A_G | TRNK | TRNA | 0.4009898 | 0.244 | MT-TK | MERRF (Myoclonic Epilepsy with Ragged Red Fibers); Other - LD / Depressive mood disorder / leukoencephalopathy / HiCM |
| 8356_T_C | TRNK | TRNA | 0.28643853 | 0.096 | MT-TK | MERRF |
| 8363_G_A | TRNK | TRNA | 0.24829501 | 0.136 | MT-TK | MICM+DEAF / MERRF / Autism / Leigh Syndrome / Ataxia+Lipomas |
| 8528_T_C | ATP8 | V | 0.05365441 | 0.1555 | MT-ATP8/6 | Infantile cardiomyopathy / hyperammonemia |
| 8851_T_C | ATP6 | V | 0.05552538 | 0.216 | MT-ATP6 | BSN / Leigh syndrome |
| 8969_G_A | ATP6 | V | 0.02154626 | 0.082 | MT-ATP6 | Mitochondrial myopathy, lactic acidosis and sideroblastic anemia (MLASA) / IgG nephropathy |
| 8993_T_C | ATP6 | V | 0.1083952 | 0.319 | MT-ATP6 | NARP / Leigh Disease / MILS / other |
| 8993_T_G | ATP6 | V | 0.1083952 | 0.136 | MT-ATP6 | NARP / Leigh Disease / MILS / other |
| 9035_T_C | ATP6 | V | 0.13096747 | 0.139 | MT-ATP6 | Ataxia syndromes |
| 9176_T_C | ATP6 | V | 0.09252218 | 0.07 | MT-ATP6 | FBSN / Leigh Disease / Spinocerebellar Ataxia |
| 9185_T_C | ATP6 | V | 0.08352948 | 0.131 | MT-ATP6 | Leigh Disease / Ataxia syndromes / NARP-like disease / Episodic weakness and Charcot-Marie-Tooth |

**Extended Data Table 2. A phenome-wide association study (PheWAS) results for association with MSS.**

See file: ukb_heteroplasmy_nature_extended_data_Table2.txt

**Extended Data Table 3. PHESANT results for association with MSS.**

See file: ukb_heteroplasmy_nature_extended_data_Table3.txt

**Extended Data Table 4. ICD-10 codes for each cause of death.**

| **Cause of death** | **ICD-10 (Version 2010)** |
| --- | --- |
| Infections | A00 – B99, L00 – L08 |
| Neoplasms | C00 – D48 |
| Cancers | C00 – C97 |
| Solid cancers | C00 – C80, C97 |
| Hematologic cancers | C81 – C96 |
| Benign neoplasms | D00 – D48 |
| Benign diseases of the blood | D50 – D89 |
| Endocrine disorders | E00 – E90 |
| Mental and behavioral disorders | F00 – F89 |
| Neurological disorders | G00 – G99 |
| Circulatory disorders | I05 – I89 |
| Respiratory disorders | J09 – J99 |
| Digestive disorders | K20 – K93 |
| Genitourinary disorders | N00 – N98 |
| COVID-19 | U07 |
| External causes, including accidents, injuries, poisoning, and drugs and biological substances | V01 – Y89 |
| Accidents | V01 – V49 |
| Intentional self-harm | X60 – X84 |

**Extended Data Table 5. ICD-10 and ICD-9 codes for each type of cancer.**

| **Type of cancer** | **ICD-10  (Version 2010)** | **ICD-9** |
| --- | --- | --- |
| Lip, oral cavity, and pharynx | C00 – C14 | 140 – 149 |
| Digestive organs | C15 – C26 | 150 – 157, 159 |
| Respiratory and intrathoracic organs (lung) | C30 – C39 | 160 – 165 |
| Bone and articular cartilage | C40, C41 | 170 |
| Skin (Malignant melanoma) | C43 | 172 |
| Mesothelial and soft tissue | C45 – C49 | 158, 171, 176 |
| Breast (in women) | C50 | 174, 175 |
| Female genital organs | C51 – C58 | 179 – 184 |
| Male genital organs | C60 – C63 | 185 – 187 |
| Urinary tract | C64 – C68 | 188, 189 |
| Eye, brain, and other parts of central nervous system | C69 – C72 | 190 – 192 |
| Thyroid and other endocrine glands | C73 – C75 | 193, 194 |
| Ill-defined, secondary, and unspecified sites | C76 - C80 | 195 – 199 |
| Lymphoid, hematopoietic, and related tissue | C81 – C96 | 200 – 208 |
| Lymphoma | C81 – C86 | 200 – 202 |
| Leukemia | C91 – C95 | 204 – 208 |

**List of Extended Data Figures**

**Extended Data Fig. 1.** Flowchart.

**Extended Data Fig. 2.** Data QC and participant exclusion criteria.

**Extended Data Fig. 3.** Hazard ratios (95% confidence intervals) for all-cause mortality by heteroplasmy count adjusted for nonsense mutation and MLC score.

**Extended Data Figures**

**Extended Data Fig. 1. Flowchart**

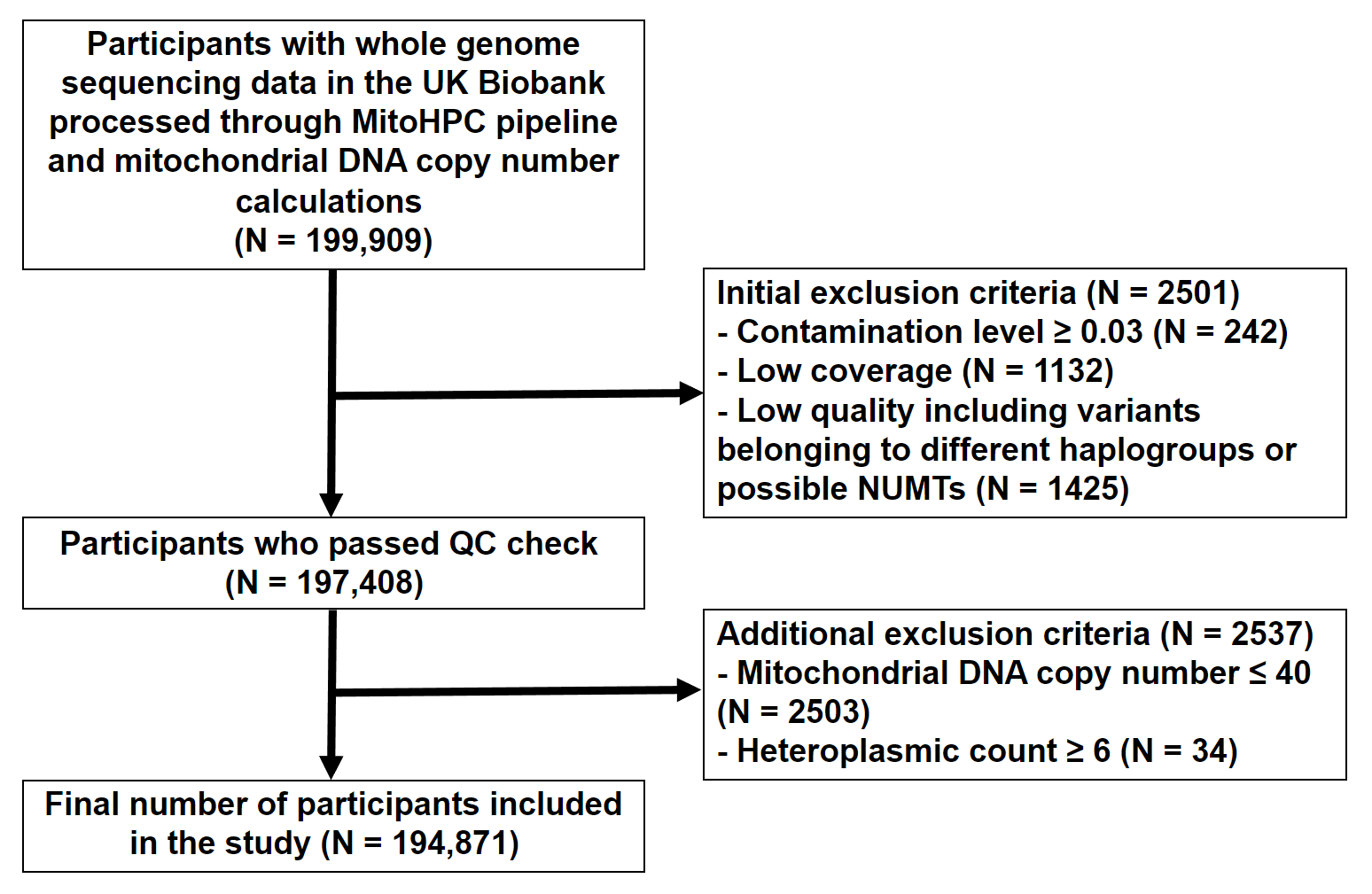

**Extended Data Fig. 1. Flowchart.** Of the 199,919 samples with outputs from MitoHPC variant calling, we calculated mtDNA copy number (mtDNA-CN) on 199,909 samples. We first excluded 2501 participants who did not meet QC criteria (potential mitochondrial contamination, low minimum base coverage, low mean base coverage, 2 or more variants belonging to a different mitochondrial haplogroup, and multiple variants predicted to be Nuclear-encoded mitochondrial sequences [NUMTs]). We further removed participants with mtDNA-CN less than 40 or participants with a heteroplasmic count above 5. 194,871 participants were included in the study for downstream analysis

**Extended Data Fig. 2.** **Data QC and participant exclusion criteria.**

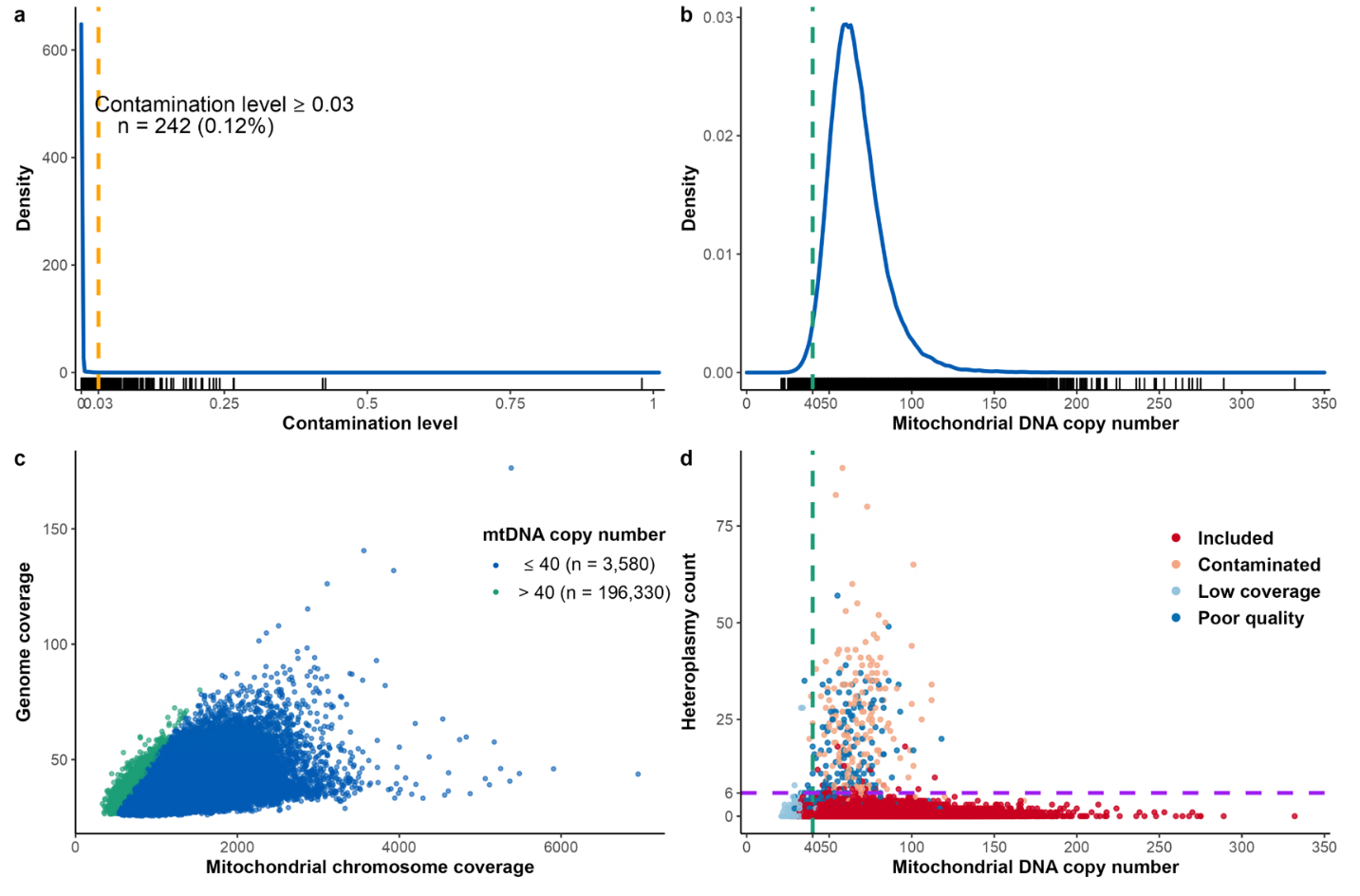

**Extended Data Fig. 2.** **Data QC and participant exclusion criteria.** (a) Contamination level for each sample (participant) plotted in a density plot. Each sample is represented by a vertical line in the rug plot along the X-axis. Orange dashed line indicates a contamination level of 3%. Samples with 3% or more contamination were excluded from analysis (n = 242). (b) Density plot of mitochondrial DNA copy number (mtDNA-CN) for all samples. Each sample is represented by a vertical line in the rug plot along the x-axis. Green dashed line indicates the cutoff (mtDNA-CN ≤ 40) for low mtDNA-CN samples (n = 3,580). (c) A scatter plot of mitochondrial chromosome coverage versus genome coverage stratified by mtDNA-CN cutoff. Samples with green dots have low mtDNA-CN and were excluded from analysis. (d) A scatter plot of mtDNA-CN and heteroplasmy count per sample in the entire dataset. Red dots are samples there were included in the study. Orange dots indicate samples that did not pass contamination threshold. Light blue dots indicate samples that were excluded due to low mitochondrial coverage defined as mean coverage less than 500 or minimum coverage less than 100. Dark blue dots indicate samples that were excluded that had other indicators of inadequate quality such as single nucleotide variants (SNVs) from a different haplogroup than the MitoHPC-identified haplogroup, or multiple SNVs matching known NUMTs (nuclear encoded mitochondrial sequences). The vertical green dashed line indicates the threshold for CN (≤ 40) at which samples were excluded from downstream analysis. The horizontal purple line indicates participants with heteroplasmy count of 6 and above. These high heteroplasmy count participants were also excluded from analysis.

**Extended Data Fig. 3. Hazard ratios (95% confidence intervals) for all-cause mortality by heteroplasmy count adjusted for mitochondrial mutation annotations.**

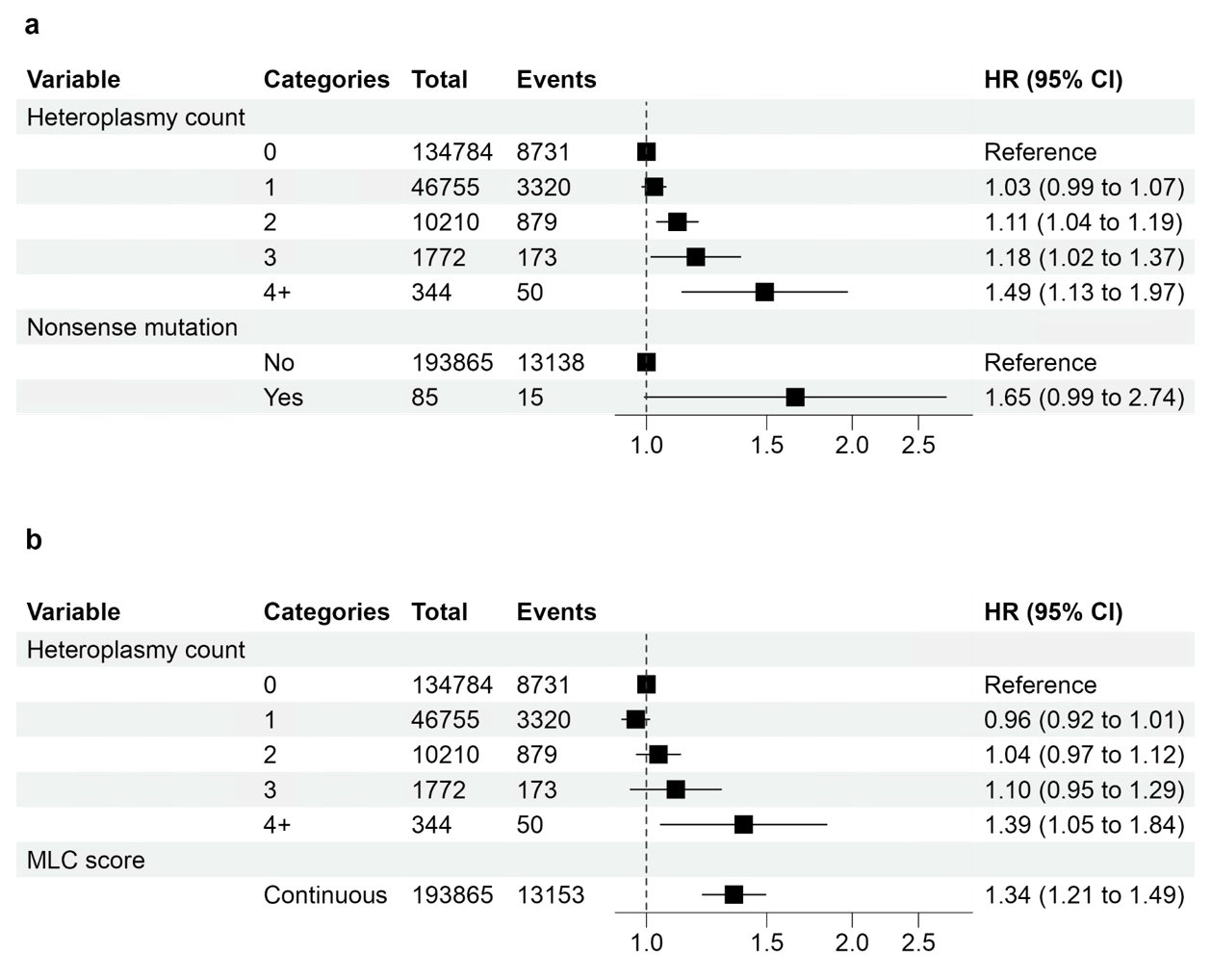

**Extended Data Fig. 3. Hazard ratios (95% confidence intervals) for all-cause mortality by heteroplasmy count adjusted for nonsense mutation and MLC score.** Hazard ratios for all-cause mortality by heteroplasmy count were adjusted for **(a)** nonsense mutation and for **(b)** MLC score. The associations were estimated from Cox proportional hazards models stratified by assessment center and adjusted for age, sex, and smoking status (never, former, or current smoker). The MLC score was generated from a randomly selected heteroplasmic SNV from each participant.
